# Reinfection with SARS-CoV-2 in the Omicron Era is Associated with Increased Risk of Post-Acute Sequelae of SARS-CoV-2 Infection: A RECOVER-EHR Cohort Study

**DOI:** 10.1101/2025.03.28.25324858

**Authors:** Bingyu Zhang, Qiong Wu, Ravi Jhaveri, Ting Zhou, Michael J. Becich, Yuriy Bisyuk, Frank Blanceró, Elizabeth A. Chrischilles, Cynthia H. Chuang, Linday G. Cowell, Daniel Fort, Carol R. Horowitz, Susan Kim, Nathalia Ladino, David M. Liebovitz, Mei Liu, Abu S. M. Mosa, Hayden T. Schwenk, Srinivasan Suresh, Bradley W. Taylor, David A. Williams, Jeffrey S. Morris, Christopher B. Forrest, Yong Chen, the RECOVER Consortium

## Abstract

**IMPORTANCE:** Post-acute sequelae of SARS-CoV-2 infection (PASC) remains a major public health challenge. While previous studies have focused on characterizing PASC and identifying its subphenotypes in children and adolescents following an initial SARS-CoV-2 infection, the risks of PASC with Omicron-variant reinfections remain unclear. Using a real-world data approach, this study investigates the risks of PASC following reinfections during the Omicron phase in the pediatric population.

**OBJECTIVE:** To investigate the risks of PASC diagnosis and 24 PASC symptoms and conditions after reinfection of SARS-CoV-2 during Omicron period in the pediatric population.

**DESIGN, SETTING, AND PARTICIPANTS:** This retrospective cohort study used data from the RECOVER consortium comprising 40 children’s hospitals and health institutions in U.S. between January 2022 and October 2023.

**EXPOSURES:** A second SARS-CoV-2 infection, confirmed by a positive polymerase-chain-reaction (PCR) or antigen tests, or a diagnose of COVID-19, occurring at least 60 days after the initial infection, compared to the initial infection.

**MAIN OUTCOMES AND MEASURES:** PASC was identified using two approaches: (1) the ICD-10- CM diagnosis code U09.9 and (2) a symptom-based definition including 24 physician-identified symptoms and conditions. Absolute risks of incident PASC were reported, and relative risks (RRs) were calculated by comparing the second infection episode with the first infection episode groups using a modified Poisson regression model, adjusting for demographic, clinical, and healthcare utilization factors through exact matching and propensity scoring matching.

**RESULTS:** A total of 465,717 individuals under 21 years old (mean [SD] age 8.17 [6.58] years; 52% male) were included. Compared to the first infection, a second infection was associated with significantly increased risk of an overall PASC diagnosis (RR, 2.08; 95% confidence interval [CI], 1.68-2.59), and with many specific conditions including: myocarditis (RR, 3.60; 95% CI, 1.46-8.86); changes in taste and smell (RR, 2.83; 95% CI, 1.41-5.67); thrombophlebitis and thromboembolism (RR, 2.28; 95% CI, 1.71-3.04); heart disease (RR, 1.96; 95% CI, 1.69 to 2.28); acute kidney injury (RR, 1.90; 95% CI, 1.38 to 2.61); fluid and electrolyte (RR, 1.89; 95% CI, 1.62 to 2.20); generalized pain (RR, 1.70; 95% CI, 1.48 to 1.95); arrhythmias (RR, 1.59; 95% CI, 1.45-1.74); abnormal liver enzyme (RR, 1.56; 95% CI, 1.24 to 1.96); fatigue and malaise (RR, 1.50; 95% CI, 1.38 to 1.64); musculoskeletal pain (RR, 1.45; 95% CI, 1.37 to 1.54); abdominal pain (RR, 1.42; 95% CI, 1.34 to 1.50); postural orthostatic tachycardia syndromes (POTS)/dysautonomia (RR, 1.35; 95% CI, 1.20 to 1.51); cognitive functions (RR, 1.32; 95% CI, 1.15 to 1.50); and respiratory signs and symptoms (RR, 1.29; 95% CI, 1.25 to 1.33). The risks were consistent across various organ systems, including cardiovascular, respiratory, gastrointestinal, neurological, and musculoskeletal systems.

**CONCLUSIONS AND RELEVANCE:** Children and adolescents face significantly higher risk of various PASC outcomes after reinfection with SARS-CoV-2. These findings suggest a cumulative risk of PASC and highlight the urgent need for targeted prevention strategies to reduce reinfections, which includes an increased emphasis on initial or re-vaccination of children.

**Key Points:** *Question:* Do children and adolescents face an increased risk of post-acute sequelae of SARS-CoV-2 infection (PASC) following reinfection during the Omicron era?

*Findings:* During the post-acute phase, children and adolescents with reinfection are at statistically significant increased risk of incident PASC outcomes, including an overall PASC diagnosis and 24 most commonly complaints/symptoms/diagnoses associated with PASC. The risks remained consistent across different demographic and clinical subgroups.

*Meaning:* These findings underscore the significant long-term health risks associated with SARS-CoV-2 reinfection in children and adolescents. Public health strategies should prioritize reinfection prevention, including enhanced vaccination efforts, to mitigate the burden of PASC in the pediatric population.

## Introduction

Since the onset of the global pandemic, post-acute sequelae of SARS-CoV-2 infection (PASC), or “Long COVID”, has emerged as a significant global concern with substantial long-term health impacts on both adults and children. The National Institutes of Health (NIH)^1^ and the Centers for Disease Control and Prevention (CDC)^2^ define PASC as new, returning, or ongoing health problems that persist for at least four weeks after initial infection, which can manifest as a wide range of symptoms and syndromes affecting multiple organ systems. Existing research has focused on the clinical features, burden, and characterization of PASC following the first infection.^3–7^ However, with the emergence of the Omicron variants, the frequency of SARS-CoV-2 reinfections has markedly increased while the acute clinical severity has generally decreased.^8–10^ This shift in the pandemic landscape has led to uncertainties regarding the risk of PASC after reinfection, particularly in the context of Omicron. Understanding the long-term consequences of reinfections is crucial, as even mild or asymptomatic cases could contribute to significant morbidity.

Several observational studies have investigated the risk of PASC after reinfection in both adults and pediatric populations.^11–21^ However, most of these studies are limited by small sample sizes, often focusing on specific populations^19,20^ or relying on survey data^13,15–18,21^, which may not be representative of the broader population. Additionally, many studies have short follow-up durations^13,15^, sometimes only a few weeks, making it difficult to capture long-term outcomes. Furthermore, the scope of PASC symptoms assessed is often narrow^14,15,17,20,21^, leaving potential manifestations unexamined. Methodological challenges, including inadequate adjustment for confounders^13–20^ and time-related biases^11^, also hinder the robustness and generalizability of findings. These limitations have left critical knowledge gaps regarding the risk and burden of PASC following reinfection during the Omicron phase.

To address these challenges, we leveraged data from the Researching COVID to Enhance Recovery (RECOVER) electronic health records (EHR) cohort, which includes comprehensive clinical data and long follow-up periods from 40 pediatric institutions. This large, diverse, and representative sample allows for a more generalizable assessment of PASC risks following reinfection. We employed robust statistical methods to minimize confounding and reduce bias. Importantly, we addressed time-related biases by carefully defining time zero for both first and second infection groups to ensure accurate risk estimation. Additionally, we conducted stratified analyses by infection severity and vaccination status to better understand potential effect modification. Using a definition commonly used in pediatric literature, we identified PASC symptoms and conditions occurring between 28 and 179 days following cohort entry.^6,22–29^ This comprehensive approach enables a thorough evaluation of the risk of PASC following reinfection compared to first infections, addressing many of the limitations identified in previous studies.

## Methods

### Data Sources and Cohort Construction

This study is part of the National Institutes of Health (NIH) funded RECOVER Initiative (https://recovercovid.org/), which aims to learn about the long-term effects of COVID-19. This study included 40 US children’s hospitals and health institutions. The EHR data was standardized to the PCORnet Common Data Model (CDM) and extracted from the RECOVER Database Version s10. More details are available in the Supplementary Materials.

We conducted a retrospective study from January 1, 2022, to October 13, 2023. We included patients under 21, who had documented SARS-CoV-2 infections after January 1, 2022 which is when Omicron variants were considered to have completely displaced earlier variants,^30^ and had at least one healthcare visit within the baseline period, defined as 24 months to 7 days before the first infection, to ensure active interaction with the healthcare system. Both the first and second infection episodes in our study occurred during the Omicron phase; for individuals in the second infection group, their first infection could have occurred before Omicron. **Figure 1** summarizes the participant selection for the first and second infection episode groups.

**Figure 1.**
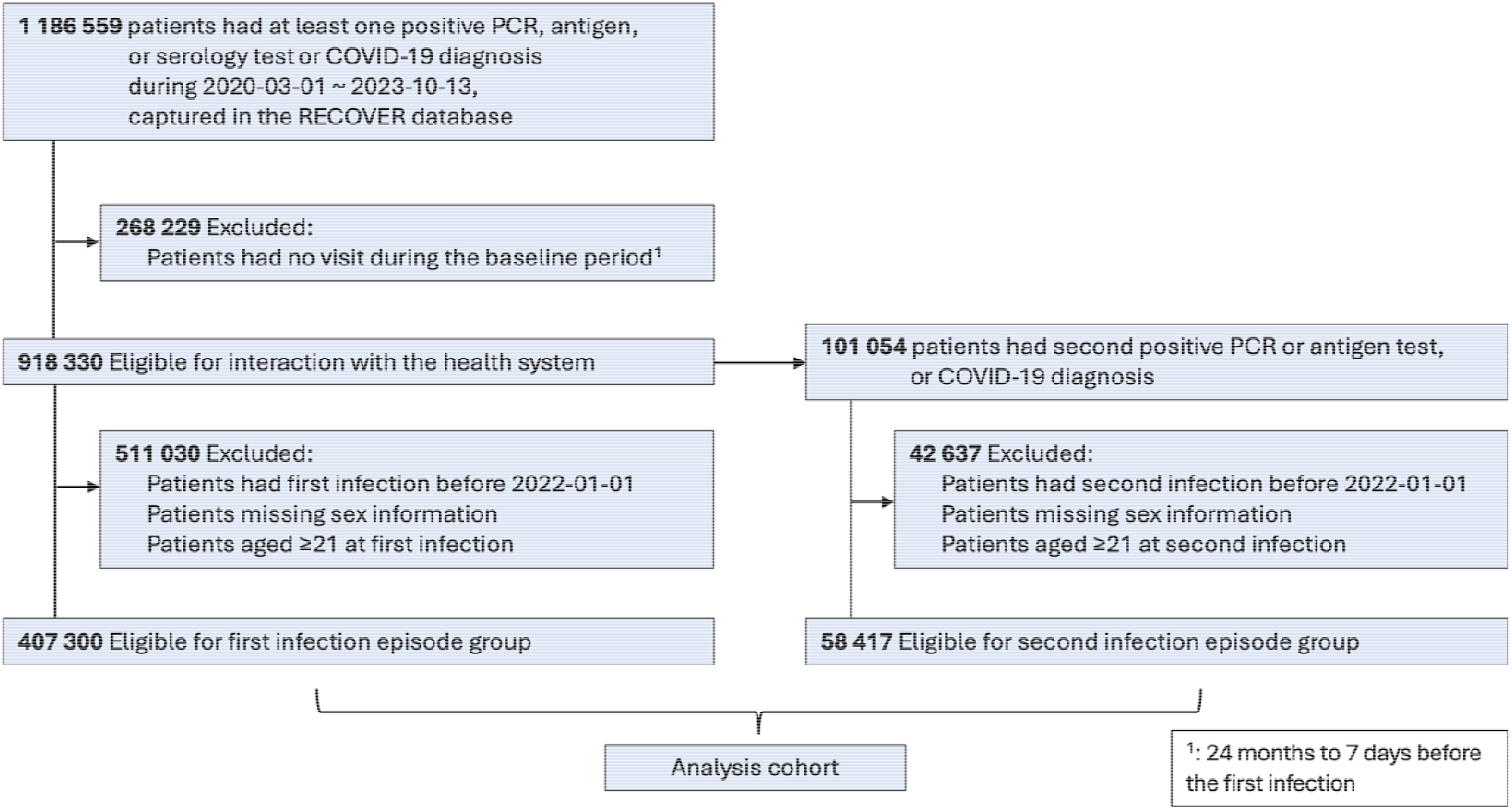
Selection of participants for first and second infection episodes.

In our study, the first documented SARS-CoV-2 infection was defined by positive polymerase-chain-reaction (PCR), serology, antigen tests, or diagnoses of COVID-19. The index date for the first infection was set as the earliest date of positive tests or COVID-19 diagnoses. The second documented SARS-CoV-2 infection was identified by positive PCR, antigen tests, or diagnoses of COVID-19 that occurred at least 60 days after their first infection. Patients whose second infection was within 60 days were put in the first infection episode group. The index date for the second infection was set as the earliest date of positive tests and COVID-19 diagnoses. Follow-up for each participant ended at the earliest of the following events: the next documented SARS-CoV-2 infection, the next documented SARS-CoV-2 infection of the matched patient, 179 days after the index date, or the end of the study period.

### PASC Outcomes and Follow-up Period

In this study, besides the International Classification of Diseases, Tenth Revision, Clinical Modification (ICD10-CM) U09.9 diagnosis code of PASC, we also included 24 symptoms and conditions identified by pediatric physicians, including abdominal pain, abnormal liver enzyme, acute kidney injury, acute respiratory distress syndrome, arrhythmias, cardiovascular signs and symptoms, changes in taste and smell, chest pain, cognitive functions, fatigue and malaise, fever and chills, fluid and electrolyte, generalized pain, hair loss, headache, heart disease, mental health disorders, musculoskeletal pain, myocarditis, myositis, postural orthostatic tachycardia syndrome (POTS) or dysautonomia, respiratory signs and symptoms, skin symptoms, and thrombophlebitis and thromboembolism.^6,22–24^

The PASC symptoms and conditions were assessed during the follow-up period, 28 to 179 days after cohort entry, in patients without a history of the specific condition during the baseline period. We used validated diagnostic codes (ICD10-CM) confirmed by board-certified pediatricians, with details of the code sets available in the Supplementary Materials.

### Covariates

A comprehensive set of patient characteristics collected during the baseline period (24 months to 7 days before the first infection) and prior to the index date were considered as confounders and adjusted through exact matching and propensity score matching to balance between groups of first and second infection episode. These included demographic factors, including age at index date, sex (female, male), and race/ethnicity (Hispanic, Non-Hispanic White (NHW), Non-Hispanic Black (NHB), Asian American/Pacific Islander (AAPI), Multiple, Other/Unknown); clinical factors, including obesity status (latest measure before index date), a chronic condition indicator defined by the Pediatric Medical Complexity Algorithm^31^ (PMCA, no chronic condition, non-complex chronic condition, complex chronic condition), and a list of pre-existing chronic conditions; health care utilization factors, including the number of inpatient visits, outpatient visits, emergency department (ED) visits, unique medications, and negative COVID-19 tests (0, 1, 2, ≥3); vaccine information, including dosage of COVID-19 vaccine before index date (0, 1, 2, ≥3) and interval since the last COVID-19 immunization (no vaccine, <4 months, ≥ 4 months); year-month of cohort entry (from January 2022 to October 2023); the severity of acute COVID-19 (“non-severe” including asymptomatic and mild, “severe” including moderate and severe); indicators from the 40 data-contributing sites.

### Statistical Analysis

We calculated the incidence of PASC outcomes in the first and second infection episodes. For each PASC symptom and condition outcome, incidence rates were calculated by dividing new diagnosis of the symptom or condition during the follow-up period by the total number of patients without any diagnoses at baseline.

To mitigate the effects of confounding, we used two matching steps, exact matching and propensity score matching, to adjust for a large number of confounders.^32,33^ We first performed exact matching using the following factors: age group (<5, 5-11, 12-20), sex, race/ethnicity, site index, vaccine doses, and cohort entry month. Within each exact-matched stratum, patients’ propensity scores were estimated by fitting a logistic regression model that includes all other covariates, including age in year. A propensity score matching was then performed within each exact-matched stratum with replacement in a 1:5 ratio. The balanced of baseline characteristics were assessed by the standardized mean difference (SMD) between the two groups, with an SMD of 0.1 or less indicating acceptable balance.^34,35^

We used the modified Poisson regression model for binary outcomes to estimate the relative risk (RR) for each outcome between comparison groups.^36^ RR is a collapsible measure, meaning the measure of association conditional on some factors remains consistent with the marginal measure collapsed over strata, which is crucial for accurate interpretation in clinical research.^37,38^

Given that vaccine status and the severity of acute COVID-19 may serve as effect modifiers, we performed stratified analyses to assess potential variations in risk across subgroups. Specifically, we stratified by the severity of acute COVID-19 (non-severe and severe) and by vaccine status (vaccinated and unvaccinated) to examine how these factors might influence the observed associations.

### Sensitivity Analysis

We conducted extensive sensitivity analyses to examine the robustness of our findings. Even though a comprehensive list of potential confounders is attempted to balance using exact and propensity score matching, residual study bias from unmeasured and systematic sources can still exist in observational studies. To address this, we performed negative control outcome experiments,^39,40^ leveraging a predefined set of 36 negative control outcomes identified by board-certified pediatricians under the assumption of a true null effect. To ensure continuity of follow-up within the healthcare system, we restricted the analysis to patients who had at least one visit during the post-acute phase (28-179 days after cohort entry). Additionally, we conducted subgroup analyses across key demographic and clinical factors, including age (0-4, 5-11, 12-20 years), sex (male and female), race/ethnicity (NHW, NHB, Hispanic, AAPI), and obesity status (obese and non-obese).

All analyses were performed using R version 4.4.0. Statistical significance was set at a p-value <0.05 (two-tailed).

## Results

### Cohort Identification

We identified an eligible total of 407,300 (87.5%) children and adolescents with the first infection episode and 58,417 (12.5%) with the second infection episode from January 1, 2022, to October 13, 2023, in the RECOVER database. Among these patients, 233,842 (50.2%) were male with a mean (SD) age of 8.17 (6.58) years.

The baseline characteristics by infection episode are detailed in **Table 1**. Among the first infection episode group, there were 170,911 Non-Hispanic White (NHW) patients (42.0%), 62,128 Non-Hispanic Black (NHB) patients (15.3%), 97,994 Hispanic patients (24.1%), 22,149 Asian American or Pacific Islander (AAPI) patients (5.4%), and others classified as multiple or unknown. In the second infection episode cohort, 26,545 patients were NHW (45.4%), 9,199 were NHB (15.7%), 14,869 were Hispanic (25.5%), 2,238 were AAPI (3.8%), and the remainder were categorized as multiple or unknown. Patients with second infection episodes tended to be older, more often females, were obese, had a higher PMCA index indicative of complex chronic conditions, and were more likely to undergo COVID-19 testing.

**Table 1.** Baseline characteristics of patients in first and second infection episodes.

|  | First infection<br>(N=407,300) | Second infection<br>(N=58,417) | Overall<br>(N=465,717) |
| --- | --- | --- | --- |
| **Age** |  |  |  |
| Median [Q1, Q3] | 7.0 [2.0, 14.0] | 9.0 [3.0, 16.0] | 7.0 [2.0, 14.0] |
| **Year of Age, no (%)** |  |  |  |
| 0 | 61718 (15.2) | 3432 (5.9) | 65150 (14.0) |
| 1 | 36876 (9.1) | 5884 (10.1) | 42760 (9.2) |
| 2 | 24836 (6.1) | 3905 (6.7) | 28741 (6.2) |
| 3 | 21058 (5.2) | 2955 (5.1) | 24013 (5.2) |
| 4 | 19822 (4.9) | 2678 (4.6) | 22500 (4.8) |
| 5 | 17116 (4.2) | 2369 (4.1) | 19485 (4.2) |
| 6 | 15795 (3.9) | 2213 (3.8) | 18008 (3.9) |
| 7 | 15401 (3.8) | 2072 (3.5) | 17473 (3.8) |
| 8 | 14680 (3.6) | 2011 (3.4) | 16691 (3.6) |
| 9 | 14708 (3.6) | 2004 (3.4) | 16712 (3.6) |
| 10 | 14482 (3.6) | 2101 (3.6) | 16583 (3.6) |
| 11 | 14712 (3.6) | 2256 (3.9) | 16968 (3.6) |
| 12 | 14161 (3.5) | 2220 (3.8) | 16381 (3.5) |
| 13 | 14907 (3.7) | 2222 (3.8) | 17129 (3.7) |
| 14 | 15482 (3.8) | 2485 (4.3) | 17967 (3.9) |
| 15 | 16099 (4.0) | 2829 (4.8) | 18928 (4.1) |
| 16 | 15972 (3.9) | 2928 (5.0) | 18900 (4.1) |
| 17 | 15841 (3.9) | 3123 (5.3) | 18964 (4.1) |
| 18 | 13827 (3.4) | 2675 (4.6) | 16502 (3.5) |
| 19 | 14585 (3.6) | 2851 (4.9) | 17436 (3.7) |
| 20 | 15222 (3.7) | 3204 (5.5) | 18426 (4.0) |
| **Sex, no (%)** |  |  |  |
| Female | 202402 (49.7) | 29473 (50.5) | 231875 (49.8) |
| Male | 204898 (50.3) | 28944 (49.5) | 233842 (50.2) |
| **Race/Ethnicity, no (%)** |  |  |  |
| NHW | 170911 (42.0) | 26545 (45.4) | 197456 (42.4) |
| NHB | 62128 (15.3) | 9199 (15.7) | 71327 (15.3) |
| Hispanic | 97994 (24.1) | 14869 (25.5) | 112863 (24.2) |
| AAPI | 22149 (5.4) | 2238 (3.8) | 24387 (5.2) |
| Multiple | 7357 (1.8) | 929 (1.6) | 8286 (1.8) |
| Other/Unknown | 46761 (11.5) | 4637 (7.9) | 51398 (11.0) |
| **Obesity, no (%)** |  |  |  |
| No | 171700 (42.2) | 21434 (36.7) | 193134 (41.5) |
| Yes | 155893 (38.3) | 24568 (42.1) | 180461 (38.7) |
| Unknown | 79707 (19.6) | 12415 (21.3) | 92122 (19.8) |
| **PMCA, no (%)** |  |  |  |
| No chronic condition | 303053 (74.4) | 37461 (64.1) | 340514 (73.1) |
| Non-complex chronic condition | 58747 (14.4) | 9486 (16.2) | 68233 (14.7) |
| Complex chronic condition | 45500 (11.2) | 11470 (19.6) | 56970 (12.2) |

| Number of negative COVID-19 tests before index date, no (%) |  |  |  |
| --- | --- | --- | --- |
| 0 | 217327 (53.4) | 21830 (37.4) | 239157 (51.4) |
| 1 | 94054 (23.1) | 13670 (23.4) | 107724 (23.1) |
| 2 | 42757 (10.5) | 8289 (14.2) | 51046 (11.0) |
| 2+ | 53162 (13.1) | 14628 (25.0) | 67790 (14.6) |
| Dosage of COVID-19 vaccine, no (%) |  |  |  |
| 0 | 318702 (78.2) | 43209 (74.0) | 361911 (77.7) |
| 1 | 15793 (3.9) | 2643 (4.5) | 18436 (4.0) |
| 2 | 55852 (13.7) | 8994 (15.4) | 64846 (13.9) |
| 2+ | 16953 (4.2) | 3571 (6.1) | 20524 (4.4) |
| Number of unique medications, no (%) |  |  |  |
| 0 | 86417 (21.2) | 9264 (15.9) | 95681 (20.5) |
| 1 | 48894 (12.0) | 5431 (9.3) | 54325 (11.7) |
| 2 | 41014 (10.1) | 4896 (8.4) | 45910 (9.9) |
| 2+ | 230975 (56.7) | 38826 (66.5) | 269801 (57.9) |

### Incidence of PASC

**Table 2** presents the number of events and absolute incidence rate per million persons per six months of PASC outcomes for the first and second infection episode groups after matching. The data indicate that second infection episode individuals generally have higher incidences. For example, the PASC diagnosis showed 903.7 and 1883.7 incidence rates per million persons per 6 months in the first and second infection groups, respectively. The highest incidence rate observed was respiratory signs and symptoms, with a rate of 61008.2 and 78342.3 in the first and second infection groups per million persons per 6 months.

**Table 2.** Number of events and incidence rate per million persons per 6 months of patients in first and second infection episodes, after exact matching and propensity score matching.

|  | Number of events |  | Incidence rate per million persons per 6 months |  |
| --- | --- | --- | --- | --- |
|  | First infection | Second infection | First infection | Second infection |
| PASC Diagnosis | 208 | 134 | 903.7 | 1883.7 |
| Acute kidney injury | 102 | 60 | 443.2 | 843.4 |
| Abdominal pain | 3700 | 1626 | 16075.4 | 22857.4 |
| Acute respiratory distress syndrome | 17 | 8 | 73.9 | 112.5 |
| Heart disease | 457 | 277 | 1985.5 | 3893.9 |
| Skin symptoms | 3726 | 1483 | 16188.3 | 20847.2 |
| Cognitive functions | 747 | 305 | 3245.5 | 4287.5 |
| Thrombophlebitis and thromboembolism | 112 | 79 | 486.6 | 1110.5 |
| Cardiovascular signs and symptoms | 1102 | 390 | 4787.9 | 5482.4 |
| Mental health | 9498 | 4046 | 41265.9 | 56876.5 |
| Arrhythmias | 1345 | 662 | 5843.6 | 9306 |
| POTS/dysautonomia | 965 | 412 | 4192.6 | 5791.7 |
| Abnormal liver enzyme | 224 | 108 | 973.2 | 1518.2 |
| Musculoskeletal pain | 3617 | 1627 | 15714.8 | 22871.5 |
| Fatigue and malaise | 1715 | 798 | 7451.2 | 11217.9 |
| Myositis | 16 | 6 | 69.5 | 84.3 |
| Myocarditis | 9 | 10 | 39.1 | 140.6 |
| Changes in the taste and smell | 17 | 15 | 73.9 | 210.9 |
| Generalized pain | 583 | 308 | 2533 | 4329.7 |
| Fluid and electrolyte | 456 | 266 | 1981.2 | 3739.3 |
| Hair loss | 211 | 66 | 916.7 | 927.8 |
| Chest pain | 1226 | 580 | 5326.6 | 8153.3 |
| Fever and chills | 4194 | 1715 | 18221.7 | 24108.6 |
| Headache | 2248 | 1023 | 9766.9 | 14380.8 |
| Respiratory signs and symptoms | 14042 | 5573 | 61008.2 | 78342.3 |

### Adjusted Relative Risk of developing PASC after Second Infections

The characteristics of the first and second infection episode groups were well-balanced after exact matching and propensity score matching (Figure S3).

**Figure 2**. demonstrates a significantly higher risk of PASC across multiple outcomes among patients with second infection episodes compared to those with first infection episodes. Notably, the overall PASC diagnosis risk was elevated (RR, 2.08; 95% confidence interval [CI], 1.68-2.59). Several specific conditions also showed increased risks, including: myocarditis (RR, 3.60; 95% CI, 1.46-8.86); changes in taste and smell (RR, 2.83; 95% CI, 1.41-5.67); thrombophlebitis and thromboembolism (RR, 2.28; 95% CI, 1.71-3.04); heart disease (RR, 1.96; 95% CI, 1.69 to 2.28); acute kidney injury (RR, 1.90; 95% CI, 1.38 to 2.61); fluid and electrolyte (RR, 1.89; 95% CI, 1.62 to 2.20); generalized pain (RR, 1.70; 95% CI, 1.48 to 1.95); arrhythmias (RR, 1.59; 95% CI, 1.45-1.74); abnormal liver enzyme (RR, 1.56; 95% CI, 1.24 to 1.96); fatigue and malaise (RR, 1.50; 95% CI, 1.38 to 1.64); musculoskeletal pain (RR, 1.45; 95% CI, 1.37 to 1.54); abdominal pain (RR, 1.42; 95% CI, 1.34 to 1.50); POTS/dysautonomia (RR, 1.35; 95% CI, 1.20 to 1.51); cognitive functions (RR, 1.32; 95% CI, 1.15 to 1.50); and respiratory signs and symptoms (RR, 1.29; 95% CI, 1.25 to 1.33).

**Figure 3**. shows that the increased risk of PASC after a second infection persist across subgroups defined by vaccination status and acute-phase severity, indicating that the heightened risk is consistent regardless of these factors. Vaccinated and unvaccinated individuals, as well as those with severe and non-severe acute COVID-19, all demonstrated higher risks following reinfection. Importantly, this analysis does not directly compare vaccinated versus unvaccinated patients or severe versus non-severe cases. Instead, it provides subgroup analyses within each group to assess whether the overall conclusion holds.

### Sensitivity Analysis

Section S3 of Supplementary Materials presents negative control outcome experiments of all PASC outcomes using 36 negative control outcomes, which indicated a slight systematic error, as shown by a minor shift in point estimates with wider CIs. Analysis restricted to patients who had at least one visit during the post-acute phase (Section S4 of Supplementary Materials) gave similar results as in the primary analyses.

**Figure 2.**
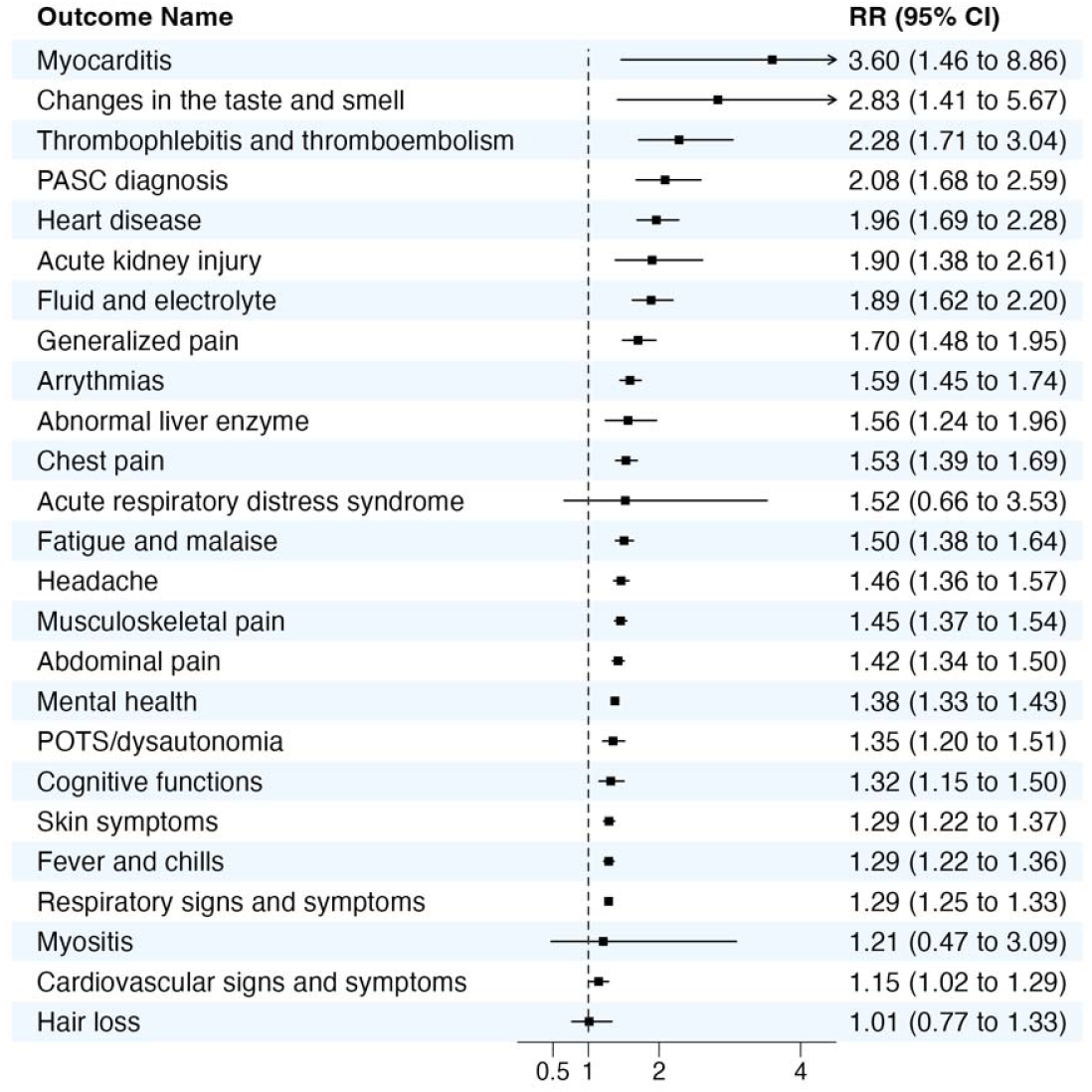
Relative Risk (RR) of incident PASC outcomes following the second infection compared with the first infection.

**Figure 3.**
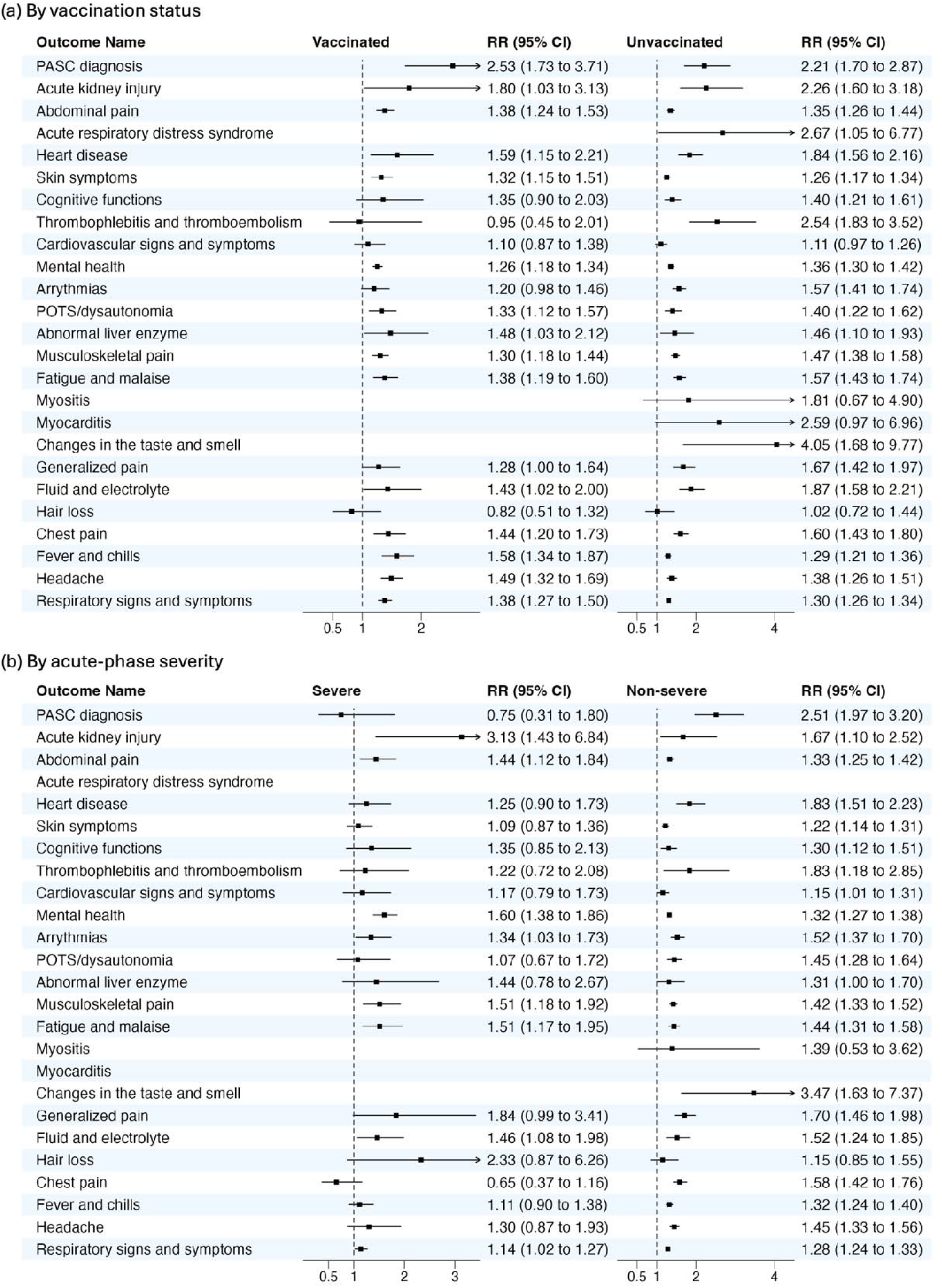
Relative Risk (RR) of incident PASC outcomes following the second infection compared with the first infection, stratified by (a) vaccine status and (b) severity of acute COVID-19.

Sections S5-8 of Supplementary Materials presents the results of a subgroup analysis, revealing a higher risk in specific populations. Adolescents over 12 displayed higher risks of PASC diagnosis, while children under 5 had higher risks of acute respiratory distress syndrome, thrombophlebitis and thromboembolism, POTS/dysautonomia, and abnormal liver enzyme (Section S5). Males showed higher risks for acute kidney injury, and females showed higher risks for PASC diagnosis, fatigue and malaise, and headache (Section S6). NHW patients exhibited the highest risk of PASC diagnosis, followed by Hispanic and NHB groups. AAPI groups showed higher risks of arrythmias (Section S7). Patients with obesity was associated with increased risk of heart disease (Section S8).

## Discussion

In this study involving 407,300 children and adolescents with a first SARS-CoV-2 infection and 58,417 with a documented reinfection, we observed an increased risk of PASC across multiple organ systems, reinforcing concerns about the long-term consequences of reinfection. These findings emphasize the ongoing risk of PASC with reinfection, regardless of severity, and suggest that the risk of PASC may be cumulative with each successive infection.

The observed increased risk of PASC in children and adolescents experiencing SARS-CoV-2 reinfection aligns with emerging immunological insights. Research has demonstrated that while T-cell memory responses remain robust and cross-reactive across various SARS-CoV-2 variants, including Omicron, B-cell responses, particularly the generation of neutralizing antibodies, may wane over time, potentially compromising immunity upon reinfection.^41^ This decline in humoral immunity could elucidate the heightened susceptibility to PASC observed in our pediatric cohort. A recent study critically illuminates the dynamic nature of immune protection, revealing that natural infection protection dramatically changed during the Omicron era, with immunity waning rapidly within a year—a pattern that may contribute to increased reinfection risks.^42^ Additionally, the Omicron variant has been associated with an increased incidence of reinfections compared to previous strains. Data indicate that the percentage of reinfections among all reported SARS-CoV-2 infections rose significantly during the Omicron BQ.1/BQ.1.1 period, particularly among younger adults.^43^ This trend underscores the necessity of evaluating reinfection risks specific to Omicron in pediatric populations, as our study suggests similar vulnerabilities in younger demographics.

The pediatric findings are further contextualized by research on long COVID in adults, which has documented cumulative symptom complexity with repeated infections, suggesting a potential mechanistic parallel across age groups. Survivor Corps research, for example, highlights how adult long COVID populations experience worsening health outcomes with successive infections, reinforcing concerns about the long-term effects of reinfection.^44^ These immunological and epidemiological observations underscore the importance of comprehensive longitudinal monitoring and targeted interventions for individuals experiencing SARS-CoV-2 reinfections, particularly among pediatric populations.

Our study addresses limitations in prior literature by employing a stronger methodological framework to account for time-related biases when assessing reinfection outcomes. For example, Bowe et al.^11^ analyzed reinfection-related risks using the US Department of Veteran Affairs’ national healthcare database and found that reinfection further increased the likelihood of sequelae across multiple organ systems. However, their study introduced a critical confounding factor, “time since infection”, by assigning time zero for the non-reinfection group based on the distribution of the calendar time of the first infection of those in the reinfection group, as PASC symptoms are more likely to occur closer to the time of the most recent infection. Our approach minimizes such biases, leading to more robust and reliable estimates of the risk associated with reinfection.

Prior research has demonstrated that vaccination reduces the risk of PASC by preventing initial infections.^24^ Since reinfection further increases the likelihood of PASC, ensuring children receive vaccines can be a key strategy in mitigating long-term health consequences. However, with the milder symptoms associated with Omicron variants and delayed availability of vaccine for children less than 5 years old, vaccination rates in children have been stagnant. Older children have not received booster doses with more recent versions of vaccine and very few younger children have received any doses of vaccine.^45^ These findings support the idea of a general “COVID fatigue” that is prevalent among the public that does not perceive COVID-19 as an ongoing threat that requires them to seek additional immunity. At the same time, the public has ongoing frustration about the lack of treatment options for PASC.^46^ With prior studies demonstrating that vaccination reduces the risk of PASC and with an increased risk of PASC with each infection, these findings could help motivate more families to seek vaccination.

Our study has several strengths. First, we leveraged the RECOVER EHR database to construct a large pediatric cohort that captured longitudinal follow-up data after the first and second SARS-COV-2 infection episodes, which provides robust statistical power and generalizability to pediatric populations in the United States. Second, we used propensity score matching which incorporated hundreds of covariates and balanced the characteristics of the two comparison groups. This allowed for better adjustment for confounders and reduced the influence of non-linear confounder effects compared to traditional regression methods.^33^ Third, we used RR as our comparative measure, which allows for accurate interpretation in clinical research.^37,38^ Lastly, we conducted an extensive sensitivity analysis including: 1) negative control experiments^39,40^ to address systematic bias and control unmeasured confounders, 2) analyses across different selected populations, and 3) a series of subgroup analyses to examine the robustness of our findings.

### Limitation

This study has several limitations that should be acknowledged. First, the documented SARS-CoV-2 infection in the EHR database is subject to potential misclassification, with asymptomatic or mild cases going unrecorded and with varying testing practices across populations. To address this, we incorporated a combination of PCR and antigen tests along with diagnosis codes for COVID-19 to more precisely define our cohort, but some underdiagnosis or misclassification may still persist. Second, differences in the healthcare utilization between the comparison groups may have influenced the observed outcomes. Patients with multiple infections may have had more frequent healthcare interactions, increasing the likelihood of diagnosis and documentation of PASC outcomes. To address this, we included healthcare utilization factors such as hospital visits and diagnostic tests as confounders in our models. Third, underrepresentation of certain racial/ethnic groups and non-obese patients may affect the generalizability of our findings. Lastly, our study did not investigate potential differences in the timing of onset, persistence, or resolution of these outcomes after the first versus the second infection.

## Conclusion

In summary, this study demonstrates a heightened risk of PASC in children and adolescents following a SARS-CoV-2 reinfection. Given that vaccination has been shown to reduce the risk of PASC, these findings underscore the importance of reinforcing public health efforts to promote vaccination among adolescents and younger children. Continued research is essential to enhance our understanding of PASC mechanisms, identify high-risk subgroups, and refine prevention and management strategies to mitigate the long-term impacts of COVID-19 in this vulnerable population.

## Data Availability

All data produced in the present study are available upon reasonable request.

## Disclosures

### Disclaimer

This content is solely the responsibility of the authors and does not necessarily represent the official views of the RECOVER Initiative, the NIH, or other funders.

## Acknowledgements

This study is part of the NIH Researching COVID to Enhance Recovery (RECOVER) Initiative, which seeks to understand, treat, and prevent the post-acute sequelae of SARS-CoV-2 infection (PASC). For more information on RECOVER, visit https://recovercovid.org/.

We would like to thank the National Community Engagement Group (NCEG), all patient, caregiver and community Representatives, and all the participants enrolled in the RECOVER Initiative. We would like to thank the patient representative Aaron Thomas Martinez for the helpful suggestions.

## Data Availability

The results reported in this study are based on detailed individual-level patient data compiled as part of the RECOVER program. Due to the high risk of reidentification based on the number of unique patterns in the date, patient privacy regulations prohibit us from releasing the data publicly. The data are maintained in a secure enclave, with access managed by the program coordinating center to remain compliant with regulatory and program requirements. Please direct requests to access the data, either for reproduction of the work reported here or for other purposes, to the RECOVER EHR Pediatric Coordinating Center.

