## Supplemental Materials for "Reinfection with SARS-CoV-2 in the Omicron Era is Associated with Increased Risk of Post-Acute Sequelae of SARS-CoV-2 Infection: A RECOVER-EHR Cohort Study"

### Section S1 Supplemental Methods

#### A. Description of electronic health records (EHR) data

The real-world data utilized in our analysis is derived from electronic health records (EHRs), covering a wide range of healthcare interaction information routinely collected and stored by hospitals. This includes clinical data such as diagnoses and treatments, laboratory and test results, and administrative data including patient demographics and billing information. The hospital-based EHR data from the Researching COVID to Enhance Recovery (RECOVER) Initiative COVID-19 Database served as the basis for defining and determining exposure, outcomes, and covariates. Unlike General Practitioner (GP) data, self-reported data, or external data sources, our study used the structured, standardized EHR entries made by healthcare providers within hospital settings. EHR data provides a more detailed and integrated view of a patient's health status, medical history, and healthcare interactions across various providers and settings.

#### B. RECOVER population and generalizability

The National Institutions of Health (NIH) launched the new RECOVER initiative in 2021 to leverage electronic health record (EHR) data to better identify and characterize patients with post-acute sequelae of SARS-CoV-2 infection (PASC). RECOVER obtains EHRs from three large national healthcare networks within the United States, covering regional catchment areas across 40 states. These networks collectively hold the EHRs of over 60 million patients, including records from more than 7 million individuals who have been affected by COVID-19. RECOVER collaborates with the National Institutes of Health's (NIH) All of Us Research Program, which contributes additional health records to this vast database. Together, these sources comprise one of the world's largest collections of EHRs.

In our study, participating institutions in this study included: Cincinnati Children’s Hospital Medical Center, Children’s Hospital of Philadelphia, Children’s Hospital of Colorado, Columbia University Irving Medical Center, Duke University, Emory, Intermountain Healthcare, University of Iowa Healthcare, University Medical Center New Orleans (LSU) – Institute for Public Health Innovation (IPHI), Ann & Robert H. Lurie Children’s Hospital of Chicago, Medical College of Wisconsin, University of Miami, University of Michigan, University of Missouri, Montefiore, Mount Sinai, Medical University of South Carolina, Children’s National Medical Center, Nationwide Children’s Hospital, Nicklaus Children’s Hospital, University of Nebraska Medical Center, Nemours Children’s Health System (in Delaware and Florida), Northwestern University, New York University School of Medicine, OCHIN, Inc., Ochsner Health System, Ohio State University, University of Pittsburgh/UPM, Pennsylvania State University, Seattle Children’s Hospital, Stanford Children’s Health, Temple University, University of California, San Francisco, University of Florida/UF Health, USF Tampa, University of Utah, UT Southwestern Medical Center, Vanderbilt University Medical Center, Wake Forest Baptist Health, and Weill Cornell Medical College. For this study, we used the s10 version of the data, collected till October 2023.

#### C. Provenance and fidelity of the data

The PASC outcomes analyzed in this study were extracted from standardized electronic health records (EHR) data provided by the RECOVER consortium, which utilizes the PCORnet Common Data Model (CDM) across 40 institutions. The PCORnet CDM ensures consistency in data formatting and structure, allowing for harmonization across different healthcare systems. All participating institutions adhered to the PCORnet CDM, which enforces standardized data collection and reporting practices. For further details, the PCORnet CDM is described in the following resources:

- <https://pcornet.org/data/>
- <https://pcornet.org/data/common-data-model/>
- <https://pcornet.org/wp-content/uploads/2023/04/PCORnet-Common-Data-Model-v61-2023_04_031.pdf>

The database version s10 refers to the specific quarterly snapshot of the dynamically updated RECOVER database used for this study. The database is refreshed quarterly to incorporate newly available data, and versioning ensures reproducibility by allowing researchers to reference the exact dataset used for their analyses.

The EHR data in this study were primarily entered by physicians and healthcare providers during routine clinical care. These entries include diagnostic codes (ICD-10-CM, ICD-10, ICD-9-CM, and SNOMED) that are widely used for billing, clinical documentation, and research purposes. All outcomes were derived directly from these validated diagnostic codes stored in the EHR systems.

To ensure the accuracy and relevance of these codes, the diagnostic code sets were reviewed and confirmed by two board-certified pediatricians (RJ and CF), who are co-authors of this manuscript. This review process ensured that the diagnostic codes used in the study were appropriate for capturing the PASC outcomes of interest.

#### D. Cohort definition and observation windows

We identified the study cohort by selecting:

- Children and adolescents had at least one positive PCR, antigen, or serology test, or COVID-19 diagnosis during 2020-03-01 to 2023-10-13
- Users of the healthcare systems, defined as having at least one primary care visit during the baseline period (either in-person, via phone, or through telehealth)
- Participants had first or second infection after 2022-01-01
- Participants aged under 21 years at cohort entry
- Participants with no previous specific PASC outcome during the baseline period when assessing the particular PASC outcome within the follow-up period

Baseline period is defined as 24 months to 7 days before the first infection, and follow-up period is defined as 28 to 179 days after cohort entry. The outcomes of interest were assessed during the follow-up period. Both the first and second infection episodes in our study occurred during the Omicron phase (after 2022-01-01). For individuals in the second infection group, their first infection could have occurred before Omicron.

**Figure S1** shows the observation windows of patients with first and second SARS-CoV-2 infections, and **Figure S2** presents the distribution of first and second SARS-CoV-2 infection from March 2020.

**Figure S1**. Observation windows of patients with first and second SARS-CoV-2 infections.

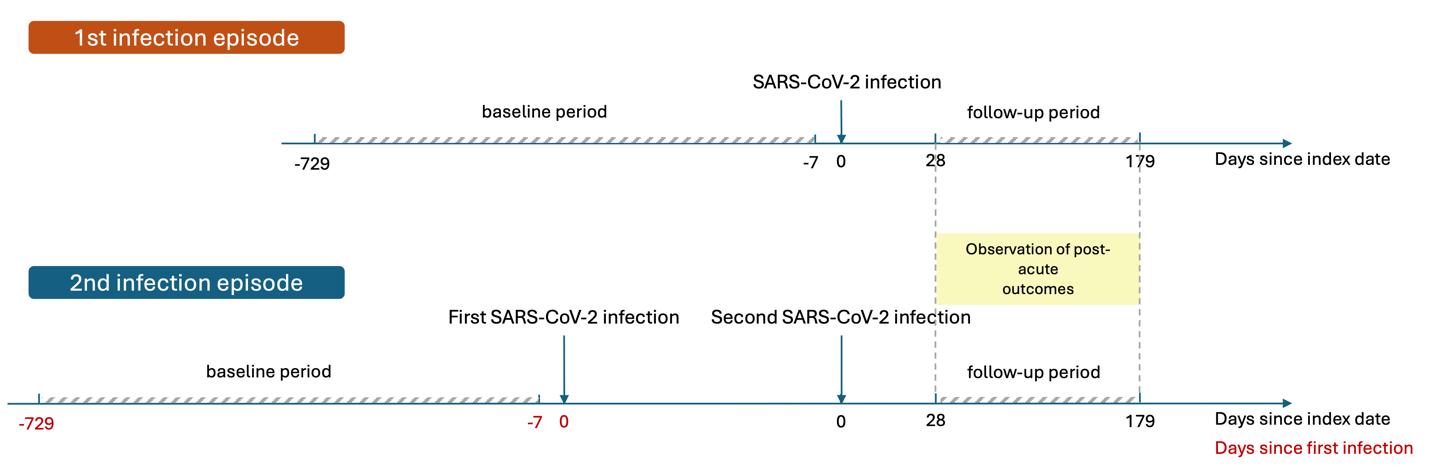

**Figure S2**. Distribution of first and second SARS-CoV-2 infection from March 2020.

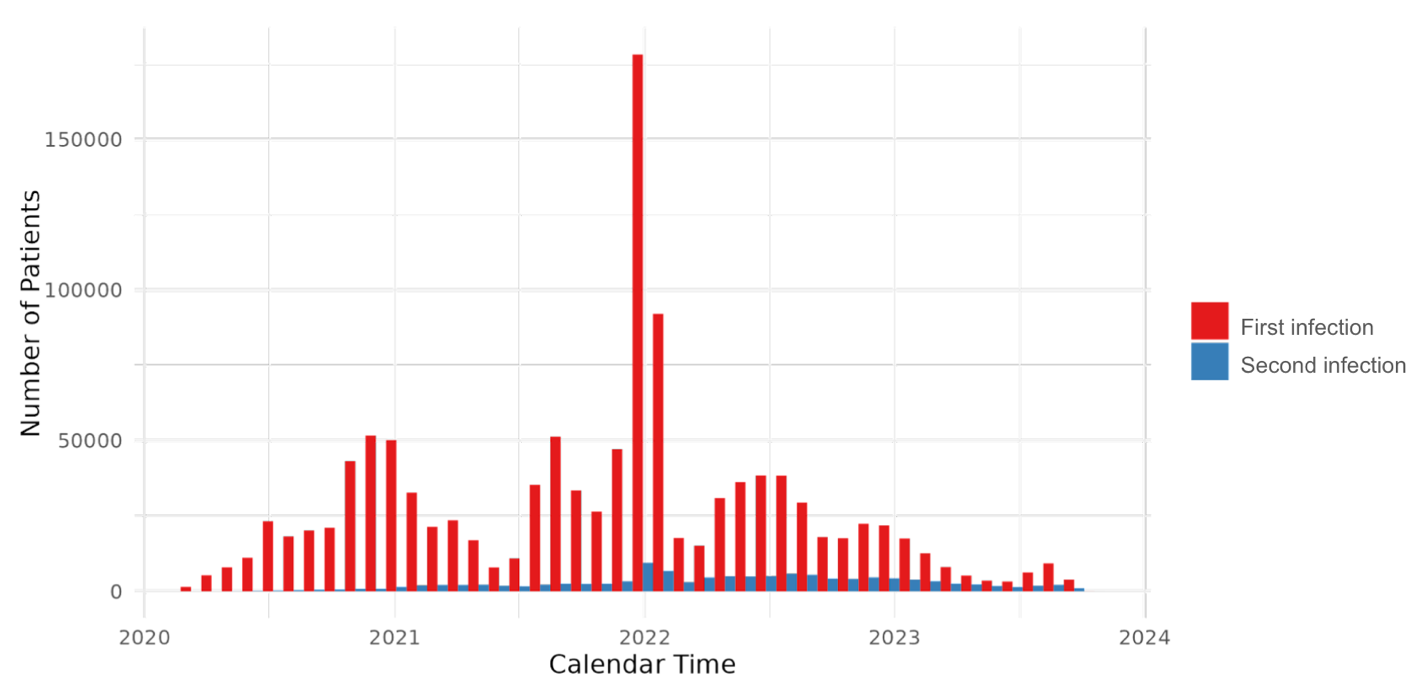

#### E. Study variables

**Table S1**. Variables used in the study evaluating the relative risk of PASC outcomes from second infection of SARS-CoV-2 compared to first infection in children and adolescents.

| Variable | Functional form | Values | Detail | Codes/references |
| --- | --- | --- | --- | --- |
| Treatment (i.e., Exposure) |  |  |  |  |
| First SARS-CoV-2 infection | Indicator | Yes/No | Based on the observation and visit occurrence domains. Defined as a polymerase-chain-reaction (PCR), serology, or antigen tests positive for COVID-19, or diagnoses of COVID-19. | See <https://github.com/PEDSnet/PASC/tree/main/observation_derivation_recover_ml_phenotype/specs> for detailed codes. |
| Second SARS-CoV-2 infection | Indicator | Yes/No | Based on the observation and visit occurrence domains. Defined as a polymerase-chain-reaction (PCR) or antigen tests positive for COVID-19, or diagnoses of COVID-19 and at least 60 days after their first infection. | See <https://github.com/PEDSnet/PASC/tree/main/observation_derivation_recover_ml_phenotype/specs> for detailed codes. |
| Outcome |  |  |  |  |
| PASC Diagnosis | Indicator | Yes/No | A set of diagnostic codes including ICD10CM, ICD10, ICD9CM, and SNOMED. | ^1–5^ |
| Acute kidney injury | Indicator | Yes/No |  |  |
| Abdominal pain | Indicator | Yes/No |  |  |
| Acute respiratory distress syndrome | Indicator | Yes/No |  |  |
| Heart disease | Indicator | Yes/No |  |  |
| Skin symptoms | Indicator | Yes/No |  |  |
| Cognitive functions | Indicator | Yes/No |  |  |
| Thrombophlebitis and thromboembolism | Indicator | Yes/No |  |  |
| Cardiovascular signs and symptoms | Indicator | Yes/No |  |  |
| Mental health | Indicator | Yes/No |  |  |
| Arrythmias | Indicator | Yes/No |  |  |
| POTS/dysautonomia | Indicator | Yes/No |  |  |
| Abnormal liver enzyme | Indicator | Yes/No |  |  |
| Musculoskeletal pain | Indicator | Yes/No |  |  |
| Fatigue and malaise | Indicator | Yes/No |  |  |
| Myositis | Indicator | Yes/No |  |  |
| Myocarditis | Indicator | Yes/No |  |  |
| Changes in the taste and smell | Indicator | Yes/No |  |  |
| Generalized pain | Indicator | Yes/No |  |  |
| Fluid and electrolyte | Indicator | Yes/No |  |  |
| Hair loss | Indicator | Yes/No |  |  |
| Chest pain | Indicator | Yes/No |  |  |
| Fever and chills | Indicator | Yes/No |  |  |
| Headache | Indicator | Yes/No |  |  |
| Confounding variables |  |  |  |  |
| Age (years) | Linear | NA | Based on records in the person domain. | Age is defined as the integer of (date – birth date)/365.25 |
| Sex | Indicator | Male/Female | Based on records in the person domain. | NA |
| Race/Ethnicity | 6 categories | NHW  NHB  Hispanic  AAPI  Multiple  Other/unknown | Based on records in the person domain. | NA |
| Obesity | 3 categories | Yes/No/Unknown | Based on records in the measurement domain.  If measured at age < 24*30.5 days, NHANES weight z score > 1.64  If measured at 24*30.5 < age <240*30.5, NHANES BMI z score > 1.64  If measured at age >= 240*30.5, BMI kg/m2 > 30 | NA |
| PMCA (Pediatric Medical Complexity Algorithm) | 3 categories | No chronic condition (PMCA = 0)  Non-complex chronic condition (PMCA = 1)  Complex chronic condition comorbidities (PMCA = 2) | Based on the condition occurrence and visit occurrence domains. | ^7^ |
| Diagnosis of each chronic condition cluster in baseline period | Indicator | Yes/No | 205 chronic condition clusters were defined based on the condition occurrence and visit occurrence domains. | ^1^ |
| Number of visits to emergency department in baseline period | 4 categories | 0/1/2/≥3 | Based on the condition occurrence and visit occurrence domains. | NA |
| Number of inpatient visits in baseline period | 4 categories | 0/1/2/≥3 | Based on the condition occurrence and visit occurrence domains, including Inpatient Hospital Stay, Emergency Department Admit to Inpatient Hospital Stay, and Observation Stay | NA |
| Number of outpatient visits in baseline period | 4 categories | 0/1/2/≥3 | Based on the condition occurrence and visit occurrence domains including Ambulatory/Outpatient Visit (With a Physician) and Interactive Telemedicine Service | NA |
| Number of unique medications in baseline period | 4 categories | 0/1/2/≥3 | Based on the drug exposure domain. | NA |
| Number of negative COVID-19 tests in baseline period | 4 categories | 0/1/2/≥3 | Based on the observation derivation recover domain | NA |
| Number of COVID-19 vaccine doses prior to the entry | 4 categories | 0/1/2/≥3 | Based on the immunization domain, using either the presence of a CVX code designating an administered or patient-reported dose or of a source value containing the terms “COVID” or “sars” in the immunization table. | CVX code: 207, 208, 210, 211, 212, 213, 217, 218 |
| Interval since the last COVID-19 immunization date | 3 categories | No vaccination  <4 months  ≥4 months |  |  |
| Other variables for eligibility criteria |  |  |  |  |
| Prior encounter in baseline period | Indicator | Yes/No | Based on the condition occurrence and visit occurrence domains. | NA |
| Follow-up encounter in follow-up period | Indicator | Yes/No | Based on the condition occurrence and visit occurrence domains. | NA |

Note: All the domains in the table above are based on the PCORnet common data model (CDM). More details are available through the following links: <https://pcornet.org/data/common-data-model/>, <https://pedsnet.org/data/pedsnet-common-data-model/>.

### Section S2 Supplemental Results: patient characteristic balance between patients with first and second SARS-CoV-2 infections

#### A. Propensity-score (PS) models and matching

We fitted a large-scale PS model for the study cohort with baseline patient characteristics including

- Demographics (age at index date; sex; race/ethnicity)
- Obesity status
- Chronic condition indicator as defined by the Pediatric Medical Complexity Algorithm (PMCA)
  - No chronic condition (PMCA = 0)
  - Non-complex chronic condition (PMCA = 1)
  - Complex chronic condition comorbidities (PMCA = 2)
- The existence of a list of 205 chronic conditions 24 months ~ 7 days in baseline period
- Healthcare utilization in baseline period categorized to 0,1,2, ≥3
  - Number of inpatient visits
  - Number of outpatient visits
  - Number of emergency department (ED) visits
  - Number of unique medications
  - Number of negative COVID-19 tests
- Vaccine information
  - Number of COVID-19 vaccine doses prior to the index, categorized to 0,1, ≥2
  - Interval since the last COVID-19 immunization date, categorized as no vaccine, <4 months, and ≥4 months
- Cohort entry date (index date) categorized to 1 month
- Healthcare system index

We exclude all covariates that occur in fewer than 0.1% of participants for computational efficiency. We first conduct exact matching using the following factors: age group (<5, 5-11, 12-20), sex, race/ethnicity, site index, vaccine doses, and cohort entry month. Within each exact-matching stratum, we performed an additional propensity score matching step with replacement in a 1:5 ratio, using a logistic regression model that included all other covariates, including age in year.

#### B. Empirical equipoise assessment

To assess the similarity across study groups, we present the preference score. This metric refines the propensity score by integrating treatment prevalence, facilitating an easily understood comparison. The preference score (F) is mathematically derived from the propensity score (S) and the treatment prevalence (P) using the following formula: ^8,9^

$$\ln\left( \frac{F}{1-F} \right)=\ln\left( \frac{S}{1-S} \right)-\ln\left( \frac{P}{1-P} \right).$$

**Figure S3** below illustrates the preference score distributions for treatment groups within each study cohort, which indicates the high comparability of these studies.

**Figure S3**. Distribution of preference scores for SARS-CoV-2 first and second infection populations. A greater convergence of these distributions indicates a higher similarity in the predicted likelihood of being secondly infected between the second infection episode (target cohort, in red) and first infection episode (comparator cohort, in blue) participants.

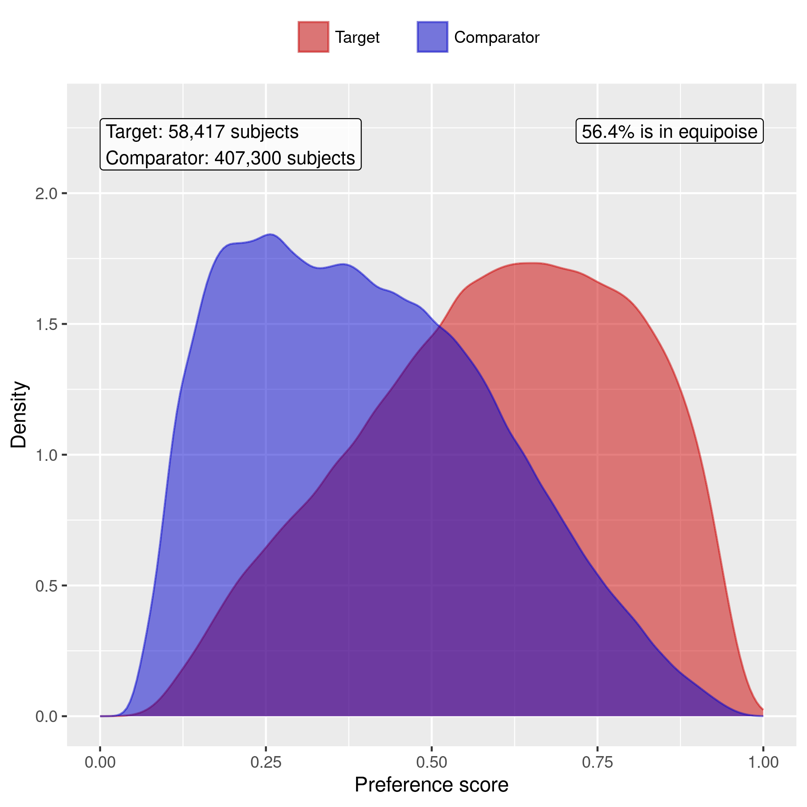

#### C. Patient characteristic balance in the primary analysis

We evaluate the balance of patient characteristics using the standardized mean difference (SMD). **Figure S4** presents the SMD of the study cohorts before and after PS score stratifications.

**Figure S4**. Patient characteristic balance before and after exact matching and large-scale PS matching. The upper panel displays the top 20 covariates with the largest SMDs before matching (red dot), while the lower panel displays the top 20 covariates with the largest SMDs after matching (blue dot).

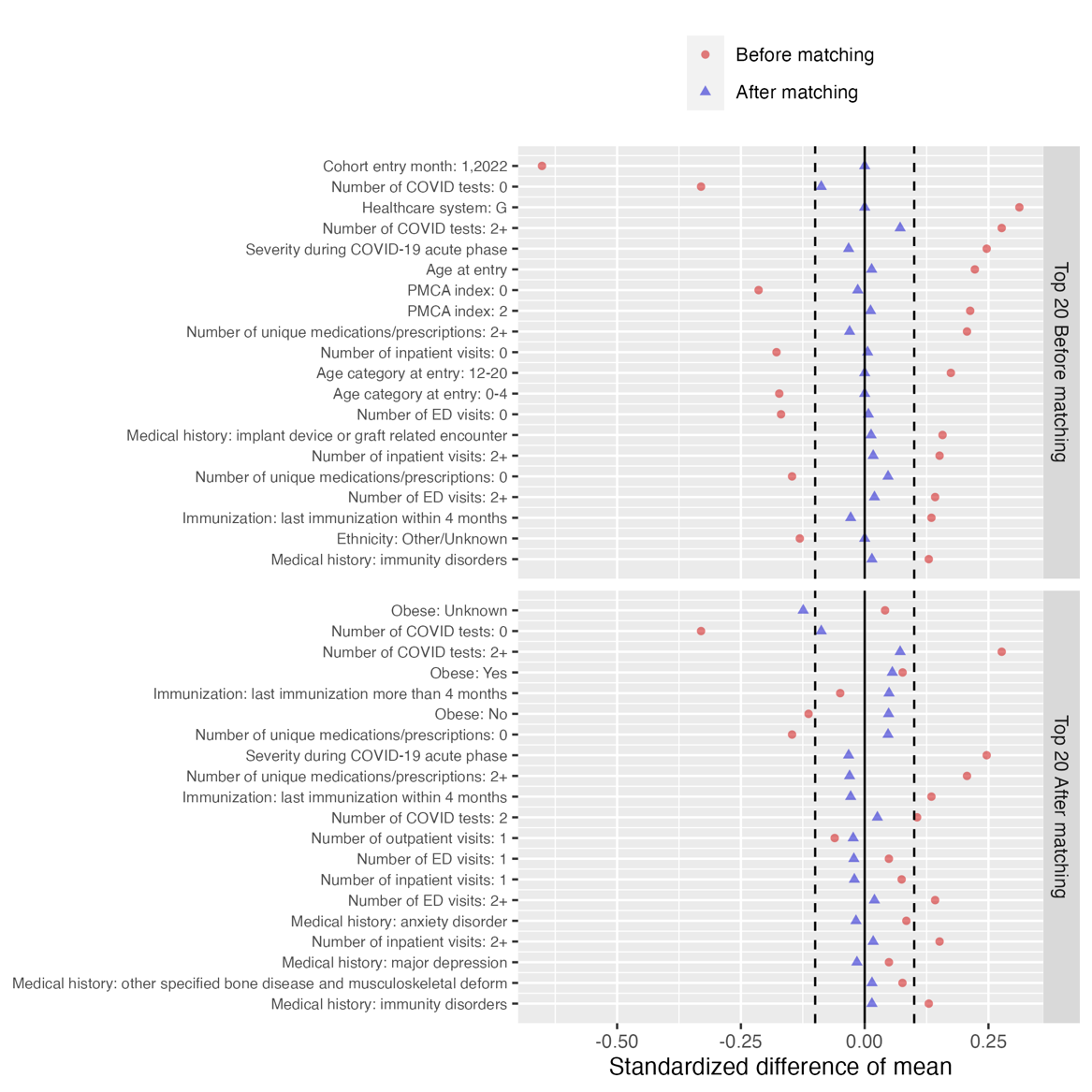

### Section S3 Supplemental Results: negative control experiments

To evaluate the robustness of our method, we conducted a list of 36 negative control outcomes to conduct the negative control experiments. Negative control outcomes, as established for our study, are defined as clinical outcomes believed to have no causal relationship to the exposure. The full list of outcomes for our research was designated by pediatric physicians. A detailed enumeration of these outcomes is presented in **Table S2**.

**Table S2**. List of negative control outcomes.

Health Conditions

Acne

Astigmatism

Autism/Autistic disorder

Closed fracture of distal end of radius

Closed injury of head

Concussion

Contact dermatitis

Diaper rash

Displacements - bone

Epilepsy

Falls

Foreign body in ear

Impetigo

Inguinal hernia

Injury of finger

Injury of free lower limb

Injury of head

Injury of left leg

Injury of right foot

Injury of right hand

Injury of right leg

Injury of upper extremity

Insect bite

Myopia

Plagiocephaly

Scoliosis

Seizure

Snoring/Obstructive sleep apnea

Speech delay

Speech dysfunction

Sprain of ankle

Tinea capitis

Tinea corporis

Tongue tie

Umbilical hernia

Wax in ear/impacted cerumen

For these 36 negative control outcomes, the null hypothesis is that the exposure—a second SARS-CoV-2 infection—exerts no influence on the outcomes. These outcomes are instrumental in detecting systematic biases in our study design and contributing to calibrating our estimated RRs.^10,11^ This list of 36 negative control outcomes were determined by two board-certified pediatricians (RJ, CF). We began by calculating the empirical null distribution for the negative control outcomes, which was then utilized to adjust the RR in the primary analysis. An in-depth discussion on the determination of the empirical null distribution and the adjustment methodology is documented in the works of Schuemie et al. ^10,11^.

To avoid the excessive uncertainty associated with the estimated causal effects of negative control outcomes, we excluded negative control outcomes with fewer than 30 events in our study cohort from our analysis. The calibrated estimates are presented in **Figure S4**.

**Figure S4**. Risks of incident PASC outcomes compared with the first infection cohort, after negative control outcomes calibration. The error bars showed the 95% confidence interval (CI) of the estimated RR.

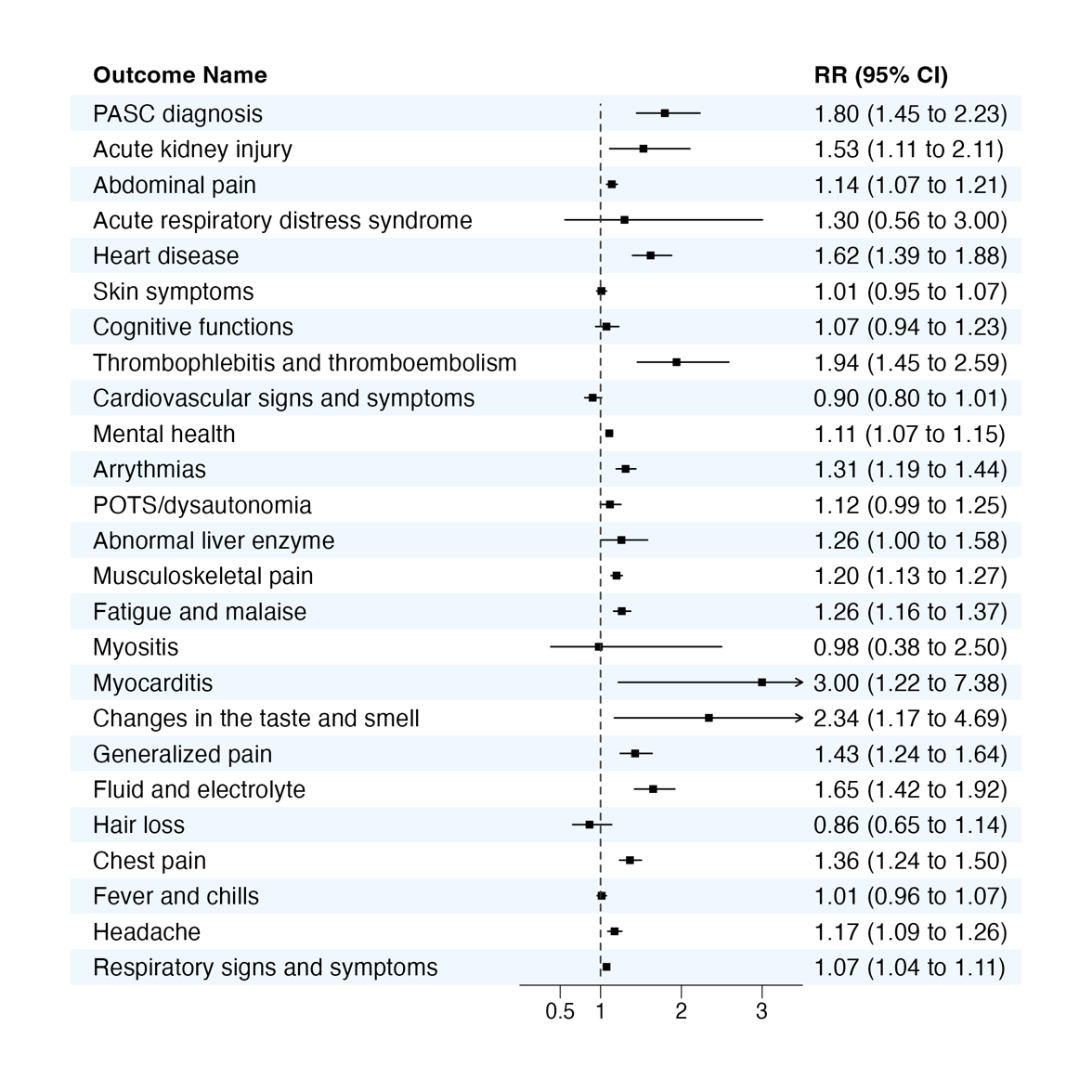

### Section S4 Supplemental Results: patients with at least one visit during post-acute phase

In the primary analysis, we do not require patients to have a follow-up visit during the post-acute phase. However, we acknowledge this could introduce informative censoring when patients were loss to follow up. We additionally restricted patients to those with at least one visit during the post-acute phase. The results are shown in **Table S3** and **Figure S5**.

**Table S3.** Number of events and incidence rate per million persons per 6 months of patients in first and second infection episodes, after exact matching and propensity score matching.

|  | Number of events | | Incidence rate per million persons per 6 months | |
| --- | --- | --- | --- | --- |
|  | First infection | Second infection | First infection | Second infection |
| PASC Diagnosis | 191 | 115 | 1171.7 | 2138.0 |
| Acute kidney injury | 109 | 61 | 781.1 | 1324.1 |
| Abdominal pain | 3711 | 1535 | 23163.5 | 29028.8 |
| Acute respiratory distress syndrome | 13 | 9 | 81.0 | 169.8 |
| Heart disease | 479 | 258 | 2939.4 | 4801.2 |
| Skin symptoms | 3729 | 1379 | 24570.9 | 27659.1 |
| Cognitive functions | 753 | 300 | 4900.1 | 5894.5 |
| Thrombophlebitis and thromboembolism | 118 | 62 | 726.2 | 1156.2 |
| Cardiovascular signs and symptoms | 1116 | 392 | 7230.9 | 7721.9 |
| Mental health | 9422 | 3795 | 59499.2 | 72255.5 |
| Arrythmias | 1338 | 615 | 8827.5 | 12267.7 |
| POTS/dysautonomia | 939 | 388 | 7917.8 | 9621.5 |
| Abnormal liver enzyme | 236 | 108 | 1507.9 | 2092.7 |
| Musculoskeletal pain | 3582 | 1497 | 23068.2 | 29156.5 |
| Fatigue and malaise | 1736 | 734 | 10741.3 | 13775.7 |
| Myositis | 22 | 9 | 151.1 | 187.5 |
| Myocarditis | 9 | 9 | 56.7 | 171.8 |
| Changes in the taste and smell | 16 | 12 | 133.3 | 299.8 |
| Generalized pain | 543 | 277 | 3962.7 | 6107.5 |
| Fluid and electrolyte | 461 | 273 | 2826.0 | 5073.8 |
| Hair loss | 199 | 61 | 1221.1 | 1136.4 |
| Chest pain | 1209 | 546 | 7745.8 | 10622.2 |
| Fever and chills | 4276 | 1621 | 44190.1 | 48956.5 |
| Headache | 2267 | 954 | 16986.5 | 21405.8 |
| Respiratory signs and symptoms | 14184 | 5327 | 87794.7 | 99979.4 |

**Figure S5**. Risks of incident PASC outcomes compared with the first infection cohort, after requiring patients having at least one visit during the follow-up period. The error bars showed the 95% confidence interval (CI) of the estimated RR.

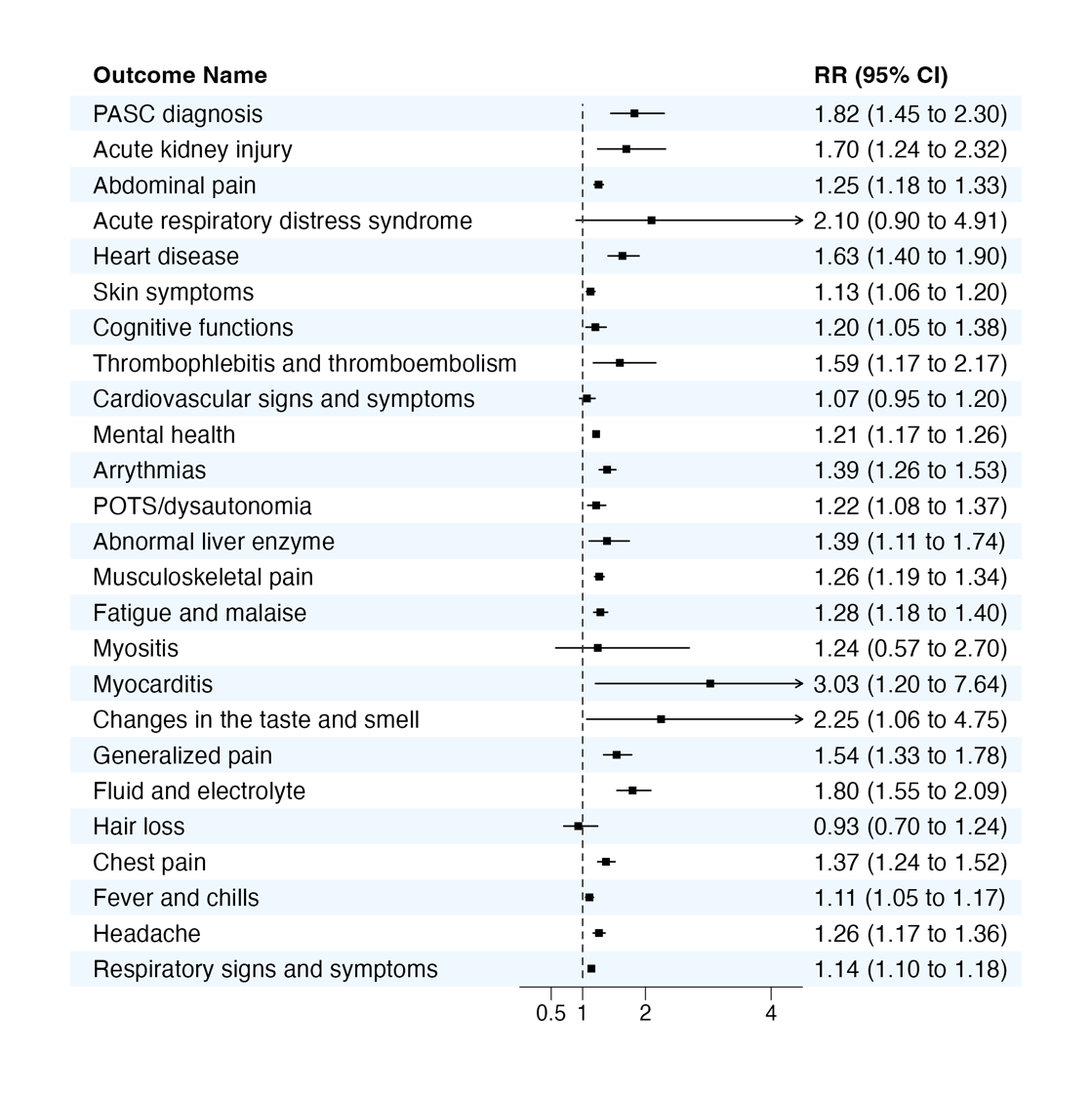

### Section S5 Supplemental Results: stratified analysis by age subgroups

We conducted subgroup analysis by age groups (0 to 4, 5 to 11, and 12 to 20 years).

**Table S4.** Number of events and incidence rate per million persons per 6 months of patients in first and second infection episodes, after exact matching and propensity score matching, age 0 to 4.

|  | Number of events | | Incidence rate per million persons per 6 months | |
| --- | --- | --- | --- | --- |
|  | First infection | Second infection | First infection | Second infection |
| PASC Diagnosis | 22 | 7 | 285.8 | 304.5 |
| Acute kidney injury | 42 | 30 | 575.7 | 1373.0 |
| Abdominal pain | 567 | 228 | 7444.3 | 10045.9 |
| Acute respiratory distress syndrome | 6 | 7 | 78.9 | 308.4 |
| Heart disease | 223 | 125 | 2908.7 | 5452.7 |
| Skin symptoms | 1893 | 683 | 25968.3 | 31606.3 |
| Cognitive functions | 517 | 217 | 7173.2 | 10077.1 |
| Thrombophlebitis and thromboembolism | 37 | 26 | 481.9 | 1133.3 |
| Cardiovascular signs and symptoms | 488 | 168 | 6382.3 | 7340.0 |
| Mental health | 1579 | 705 | 21268.9 | 31722.1 |
| Arrythmias | 411 | 222 | 5564.8 | 10036.2 |
| POTS/dysautonomia | 22 | 16 | 442.9 | 1043.1 |
| Abnormal liver enzyme | 60 | 37 | 810.9 | 1683.6 |
| Musculoskeletal pain | 399 | 160 | 5235.7 | 7042.2 |
| Fatigue and malaise | 405 | 189 | 5280.8 | 8246.7 |
| Myositis | <6 | <6 | - | - |
| Myocarditis | <6 | <6 | - | - |
| Changes in the taste and smell | <6 | <6 | - | - |
| Generalized pain | 57 | 29 | 767.0 | 1305.4 |
| Fluid and electrolyte | 230 | 144 | 2989.9 | 6267.4 |
| Hair loss | 42 | 14 | 546.2 | 609.5 |
| Chest pain | 85 | 38 | 1112.9 | 1666.0 |
| Fever and chills | 2497 | 1034 | 59390.1 | 79582.3 |
| Headache | 106 | 49 | 1701.6 | 2612.2 |
| Respiratory signs and symptoms | 7488 | 2891 | 98215.4 | 127232.0 |

**Table S5.** Number of events and incidence rate per million persons per 6 months of patients in first and second infection episodes, after exact matching and propensity score matching, age 5 to 11.

|  | Number of events | | Incidence rate per million persons per 6 months | |
| --- | --- | --- | --- | --- |
|  | First infection | Second infection | First infection | Second infection |
| PASC Diagnosis | 47 | 27 | 811.5 | 1490.2 |
| Acute kidney injury | 23 | 16 | 474.0 | 1052.5 |
| Abdominal pain | 1160 | 432 | 20389.0 | 24169.4 |
| Acute respiratory distress syndrome | <6 | <6 | - | - |
| Heart disease | 81 | 43 | 1403.1 | 2379.2 |
| Skin symptoms | 711 | 312 | 12965.0 | 18090.6 |
| Cognitive functions | 122 | 46 | 2209.8 | 2654.4 |
| Thrombophlebitis and thromboembolism | 22 | 8 | 380.7 | 442.1 |
| Cardiovascular signs and symptoms | 255 | 97 | 4643.0 | 5637.5 |
| Mental health | 1963 | 857 | 34648.4 | 48399.9 |
| Arrythmias | 218 | 87 | 3973.4 | 5063.3 |
| POTS/dysautonomia | 112 | 50 | 2698.2 | 3802.6 |
| Abnormal liver enzyme | 45 | 21 | 804.6 | 1196.1 |
| Musculoskeletal pain | 837 | 408 | 14895.7 | 23168.7 |
| Fatigue and malaise | 294 | 148 | 5121.7 | 8220.1 |
| Myositis | <6 | <6 | - | - |
| Myocarditis | <6 | <6 | - | - |
| Changes in the taste and smell | <6 | <6 | - | - |
| Generalized pain | 150 | 74 | 2989.5 | 4687.0 |
| Fluid and electrolyte | 77 | 46 | 1325.9 | 2531.9 |
| Hair loss | 38 | 20 | 656.7 | 1104.6 |
| Chest pain | 344 | 153 | 6030.8 | 8559.7 |
| Fever and chills | 842 | 342 | 24052.8 | 30954.2 |
| Headache | 637 | 246 | 13011.1 | 15973.5 |
| Respiratory signs and symptoms | 3205 | 1308 | 55840.7 | 72763.5 |

**Table S6.** Number of events and incidence rate per million persons per 6 months of patients in first and second infection episodes, after exact matching and propensity score matching, age 12 to 20.

|  | Number of events | | Incidence rate per million persons per 6 months | |
| --- | --- | --- | --- | --- |
|  | First infection | Second infection | First infection | Second infection |
| PASC Diagnosis | 126 | 98 | 1393.0 | 3346.7 |
| Acute kidney injury | 40 | 30 | 537.9 | 1252.2 |
| Abdominal pain | 2017 | 946 | 22654.3 | 32798.1 |
| Acute respiratory distress syndrome | <6 | <6 | - | - |
| Heart disease | 182 | 114 | 2015.3 | 3897.8 |
| Skin symptoms | 1041 | 426 | 12358.7 | 15677.6 |
| Cognitive functions | 82 | 31 | 942.2 | 1097.5 |
| Thrombophlebitis and thromboembolism | 65 | 29 | 722.4 | 995.3 |
| Cardiovascular signs and symptoms | 345 | 123 | 4195.5 | 4611.2 |
| Mental health | 6033 | 2413 | 67216.8 | 83100.9 |
| Arrythmias | 699 | 325 | 8530.8 | 12142.8 |
| POTS/dysautonomia | 818 | 342 | 10316.5 | 13265.8 |
| Abnormal liver enzyme | 131 | 56 | 1477.6 | 1953.3 |
| Musculoskeletal pain | 2355 | 1044 | 28207.7 | 38349.2 |
| Fatigue and malaise | 1017 | 453 | 11346.7 | 15602.8 |
| Myositis | 6 | <6 | 80.0 | - |
| Myocarditis | <6 | 7 | - | 242.8 |
| Changes in the taste and smell | 11 | 9 | 178.7 | 447.0 |
| Generalized pain | 397 | 194 | 5902.4 | 8792.3 |
| Fluid and electrolyte | 177 | 100 | 1962.8 | 3419.2 |
| Hair loss | 129 | 31 | 1429.7 | 1059.5 |
| Chest pain | 791 | 373 | 9487.1 | 13756.3 |
| Fever and chills | 560 | 271 | 8391.5 | 12389.4 |
| Headache | 1576 | 736 | 19878.0 | 28542.5 |
| Respiratory signs and symptoms | 2857 | 1269 | 31736.2 | 43502.1 |

**Figure S6**. Risks of incident PASC outcomes compared with the first infection cohort, by age subgroup. The error bars showed the 95% confidence interval (CI) of the estimated RR.

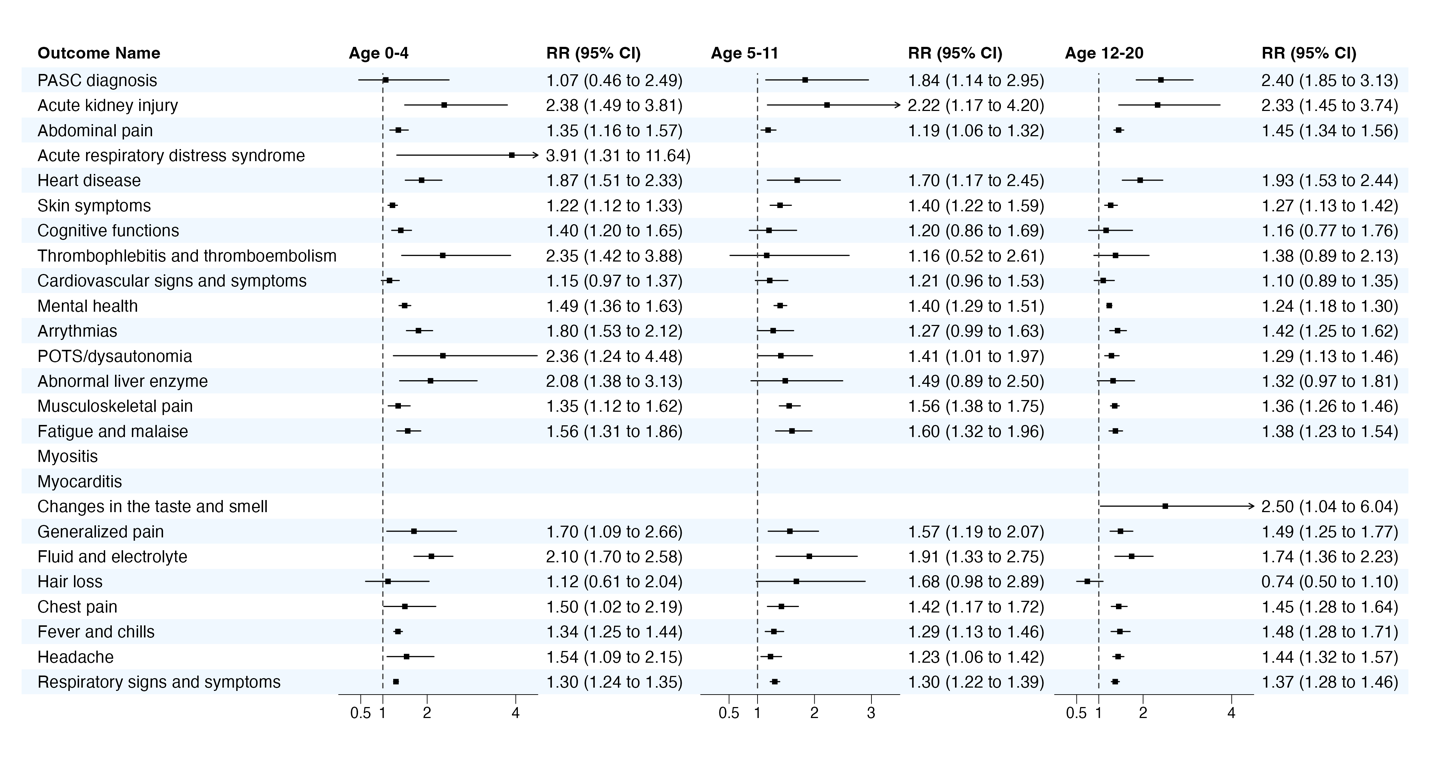

### Section S6 Supplemental Results: stratified analysis by sex subgroups

We conducted subgroup analysis by sex.

**Table S7.** Number of events and incidence rate per million persons per 6 months of patients in first and second infection episodes, after exact matching and propensity score matching, female.

|  | Number of events | | Incidence rate per million persons per 6 months | |
| --- | --- | --- | --- | --- |
|  | First infection | Second infection | First infection | Second infection |
| PASC Diagnosis | 111 | 84 | 958.6 | 2339.3 |
| Acute kidney injury | 50 | 29 | 514.4 | 961.3 |
| Abdominal pain | 2336 | 1056 | 20530.1 | 29857.7 |
| Acute respiratory distress syndrome | <6 | <6 | - | - |
| Heart disease | 215 | 111 | 1856.5 | 3092.2 |
| Skin symptoms | 1889 | 777 | 17414.9 | 23148.4 |
| Cognitive functions | 271 | 117 | 2450.7 | 3396.4 |
| Thrombophlebitis and thromboembolism | 67 | 41 | 582.0 | 1148.9 |
| Cardiovascular signs and symptoms | 518 | 185 | 4761.3 | 5499.6 |
| Mental health | 5532 | 2344 | 48756.6 | 66435.0 |
| Arrythmias | 771 | 374 | 7135.4 | 11122.9 |
| POTS/dysautonomia | 700 | 294 | 7844.5 | 10449.0 |
| Abnormal liver enzyme | 103 | 47 | 915.7 | 1345.1 |
| Musculoskeletal pain | 2039 | 930 | 18538.2 | 27233.1 |
| Fatigue and malaise | 972 | 480 | 8466.6 | 13458.1 |
| Myositis | 6 | <6 | 58.6 | - |
| Myocarditis | <6 | <6 | - | - |
| Changes in the taste and smell | 8 | 10 | 91.7 | 367.5 |
| Generalized pain | 348 | 179 | 3607.1 | 5996.9 |
| Fluid and electrolyte | 231 | 141 | 1995.5 | 3930.8 |
| Hair loss | 129 | 50 | 1115.7 | 1395.0 |
| Chest pain | 698 | 325 | 6330.8 | 9559.0 |
| Fever and chills | 1968 | 790 | 25789.7 | 32590.2 |
| Headache | 1416 | 684 | 14540.2 | 22427.0 |
| Respiratory signs and symptoms | 6382 | 2588 | 55529.8 | 72644.3 |

**Table S8.** Number of events and incidence rate per million persons per 6 months of patients in first and second infection episodes, after exact matching and propensity score matching, male.

|  | Number of events | | Incidence rate per million persons per 6 months | |
| --- | --- | --- | --- | --- |
|  | First infection | Second infection | First infection | Second infection |
| PASC Diagnosis | 73 | 46 | 640.0 | 1309.4 |
| Acute kidney injury | 48 | 37 | 468.5 | 1173.4 |
| Abdominal pain | 1454 | 567 | 12929.2 | 16348.1 |
| Acute respiratory distress syndrome | <6 | <6 | - | - |
| Heart disease | 264 | 152 | 2316.1 | 4330.2 |
| Skin symptoms | 1861 | 699 | 17365.9 | 21164.4 |
| Cognitive functions | 510 | 198 | 4703.0 | 5927.9 |
| Thrombophlebitis and thromboembolism | 58 | 31 | 509.0 | 883.1 |
| Cardiovascular signs and symptoms | 611 | 212 | 5575.2 | 6296.2 |
| Mental health | 3990 | 1641 | 35964.1 | 47843.3 |
| Arrythmias | 614 | 271 | 5674.7 | 8134.4 |
| POTS/dysautonomia | 244 | 121 | 2865.6 | 4479.4 |
| Abnormal liver enzyme | 136 | 55 | 1238.4 | 1620.1 |
| Musculoskeletal pain | 1581 | 680 | 14306.1 | 19946.5 |
| Fatigue and malaise | 761 | 313 | 6666.9 | 8906.5 |
| Myositis | 11 | <6 | 104.8 | - |
| Myocarditis | 6 | <6 | 53.9 | - |
| Changes in the taste and smell | 7 | <6 | 77.5 | - |
| Generalized pain | 228 | 116 | 2297.3 | 3771.8 |
| Fluid and electrolyte | 237 | 139 | 2075.8 | 3956.7 |
| Hair loss | 51 | 15 | 448.4 | 428.0 |
| Chest pain | 524 | 244 | 4705.1 | 7095.3 |
| Fever and chills | 2264 | 919 | 31924.8 | 40757.4 |
| Headache | 851 | 318 | 8847.0 | 10633.5 |
| Respiratory signs and symptoms | 7633 | 2966 | 67382.1 | 84978.0 |

**Figure S7**. Risks of incident PASC outcomes compared with the first infection cohort, by sex subgroup. The error bars showed the 95% confidence interval (CI) of the estimated RR.

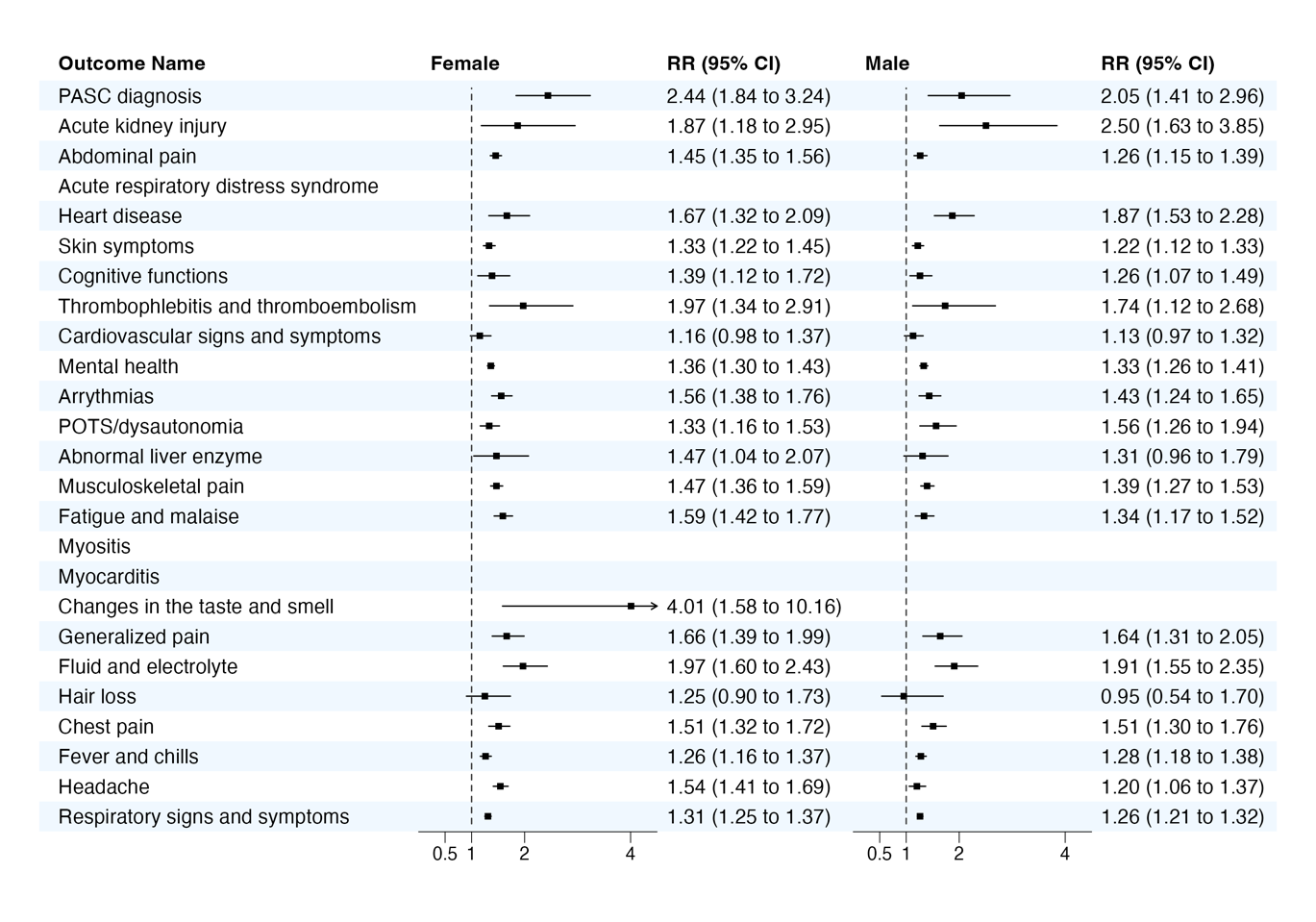

### Section S7 Supplemental Results: stratified analysis by race/ethnicity subgroups

We conducted subgroup analysis by race/ethnicity, where we included Non-Hispanic White (NHW), Hispanic, Non-Hispanic Black (NHB), and Asian American or Pacific Islander (AAPI) patients. Other race/ethnicity were not included in this analysis due to limited sample size.

**Table S9.** Number of events and incidence rate per million persons per 6 months of patients in first and second infection episodes, after exact matching and propensity score matching, NHW.

|  | Number of events | | Incidence rate per million persons per 6 months | |
| --- | --- | --- | --- | --- |
|  | First infection | Second infection | First infection | Second infection |
| PASC Diagnosis | 130 | 82 | 1103.7 | 2329.6 |
| Acute kidney injury | 79 | 43 | 766.6 | 1407.9 |
| Abdominal pain | 2058 | 900 | 17731.9 | 25881.0 |
| Acute respiratory distress syndrome | <6 | <6 | - | - |
| Heart disease | 281 | 167 | 2386.9 | 4740.9 |
| Skin symptoms | 1895 | 722 | 17037.2 | 21924.6 |
| Cognitive functions | 326 | 115 | 2866.3 | 3377.4 |
| Thrombophlebitis and thromboembolism | 68 | 44 | 580.3 | 1255.2 |
| Cardiovascular signs and symptoms | 491 | 169 | 4346.8 | 5033.1 |
| Mental health | 5787 | 2353 | 49971.6 | 67692.2 |
| Arrythmias | 808 | 359 | 7408.0 | 11007.8 |
| POTS/dysautonomia | 607 | 285 | 6623.7 | 10148.3 |
| Abnormal liver enzyme | 107 | 50 | 932.5 | 1464.7 |
| Musculoskeletal pain | 2051 | 943 | 18221.2 | 28008.6 |
| Fatigue and malaise | 1220 | 540 | 10406.4 | 15433.5 |
| Myositis | 7 | <6 | 66.1 | - |
| Myocarditis | 6 | 8 | 51.8 | 231.8 |
| Changes in the taste and smell | 11 | 9 | 123.2 | 335.2 |
| Generalized pain | 340 | 178 | 3448.0 | 6036.4 |
| Fluid and electrolyte | 277 | 138 | 2354.8 | 3918.7 |
| Hair loss | 81 | 25 | 689.9 | 711.0 |
| Chest pain | 563 | 271 | 4991.9 | 8038.8 |
| Fever and chills | 2044 | 814 | 25998.4 | 33666.2 |
| Headache | 1298 | 563 | 12824.2 | 18543.0 |
| Respiratory signs and symptoms | 6453 | 2418 | 55115.1 | 69091.8 |

**Table S10.** Number of events and incidence rate per million persons per 6 months of patients in first and second infection episodes, after exact matching and propensity score matching, Hispanic.

|  | Number of events | | Incidence rate per million persons per 6 months | |
| --- | --- | --- | --- | --- |
|  | First infection | Second infection | First infection | Second infection |
| PASC Diagnosis | 34 | 19 | 603.4 | 1093.5 |
| Acute kidney injury | 22 | 17 | 452.8 | 1130.6 |
| Abdominal pain | 1014 | 412 | 18261.8 | 24161.1 |
| Acute respiratory distress syndrome | <6 | <6 | - | - |
| Heart disease | 102 | 56 | 1813.1 | 3231.8 |
| Skin symptoms | 1035 | 381 | 19500.3 | 23478.7 |
| Cognitive functions | 214 | 93 | 3991.2 | 5615.6 |
| Thrombophlebitis and thromboembolism | 22 | 10 | 391.1 | 576.9 |
| Cardiovascular signs and symptoms | 367 | 105 | 6928.1 | 6440.8 |
| Mental health | 2028 | 902 | 36790.4 | 53207.0 |
| Arrythmias | 320 | 129 | 5894.4 | 7743.9 |
| POTS/dysautonomia | 204 | 71 | 4849.5 | 5442.6 |
| Abnormal liver enzyme | 73 | 39 | 1333.4 | 2319.1 |
| Musculoskeletal pain | 762 | 342 | 14079.0 | 20352.7 |
| Fatigue and malaise | 288 | 120 | 5158.6 | 6970.0 |
| Myositis | <6 | <6 | - | - |
| Myocarditis | <6 | <6 | - | - |
| Changes in the taste and smell | <6 | <6 | - | - |
| Generalized pain | 134 | 68 | 2709.8 | 4422.2 |
| Fluid and electrolyte | 108 | 64 | 1921.4 | 3686.4 |
| Hair loss | 67 | 23 | 1188.0 | 1323.1 |
| Chest pain | 398 | 165 | 7289.2 | 9813.3 |
| Fever and chills | 1201 | 438 | 33403.6 | 38806.6 |
| Headache | 568 | 242 | 11898.3 | 16346.0 |
| Respiratory signs and symptoms | 3986 | 1581 | 71157.4 | 91664.3 |

**Table S11.** Number of events and incidence rate per million persons per 6 months of patients in first and second infection episodes, after exact matching and propensity score matching, NHB.

|  | Number of events | | Incidence rate per million persons per 6 months | |
| --- | --- | --- | --- | --- |
|  | First infection | Second infection | First infection | Second infection |
| PASC Diagnosis | 22 | 10 | 646.1 | 930.8 |
| Acute kidney injury | 12 | 11 | 407.2 | 1180.6 |
| Abdominal pain | 488 | 215 | 14505.4 | 20174.2 |
| Acute respiratory distress syndrome | <6 | <6 | - | - |
| Heart disease | 89 | 46 | 2614.0 | 4293.5 |
| Skin symptoms | 516 | 199 | 16242.6 | 20033.8 |
| Cognitive functions | 125 | 62 | 3950.3 | 6176.9 |
| Thrombophlebitis and thromboembolism | 20 | 13 | 593.0 | 1222.1 |
| Cardiovascular signs and symptoms | 155 | 70 | 4908.9 | 6991.5 |
| Mental health | 1096 | 519 | 33098.6 | 49641.0 |
| Arrythmias | 164 | 108 | 5042.6 | 10481.7 |
| POTS/dysautonomia | 95 | 57 | 3763.6 | 7020.9 |
| Abnormal liver enzyme | 25 | 11 | 766.6 | 1064.9 |
| Musculoskeletal pain | 572 | 261 | 17563.6 | 25344.4 |
| Fatigue and malaise | 184 | 103 | 5465.2 | 9661.5 |
| Myositis | <6 | <6 | - | - |
| Myocarditis | <6 | <6 | - | - |
| Changes in the taste and smell | <6 | <6 | - | - |
| Generalized pain | 78 | 43 | 2738.1 | 4749.4 |
| Fluid and electrolyte | 74 | 51 | 2175.9 | 4751.8 |
| Hair loss | 28 | 13 | 821.7 | 1208.1 |
| Chest pain | 183 | 87 | 5578.8 | 8360.0 |
| Fever and chills | 490 | 245 | 24925.5 | 38467.1 |
| Headache | 334 | 157 | 11824.8 | 17580.4 |
| Respiratory signs and symptoms | 2240 | 935 | 66281.3 | 87667.8 |

**Table S12.** Number of events and incidence rate per million persons per 6 months of patients in first and second infection episodes, after exact matching and propensity score matching, AAPI.

|  | Number of events | | Incidence rate per million persons per 6 months | |
| --- | --- | --- | --- | --- |
|  | First infection | Second infection | First infection | Second infection |
| PASC Diagnosis | <6 | <6 | - | - |
| Acute kidney injury | <6 | <6 | - | - |
| Abdominal pain | 49 | 30 | 10044.3 | 16507.0 |
| Acute respiratory distress syndrome | <6 | <6 | - | - |
| Heart disease | <6 | <6 | - | - |
| Skin symptoms | 81 | 37 | 17945.8 | 21486.7 |
| Cognitive functions | 19 | 6 | 4041.7 | 3409.6 |
| Thrombophlebitis and thromboembolism | <6 | <6 | - | - |
| Cardiovascular signs and symptoms | 26 | 6 | 5462.5 | 3365.1 |
| Mental health | 121 | 57 | 25314.3 | 31587.1 |
| Arrythmias | 18 | 17 | 3852.3 | 9681.9 |
| POTS/dysautonomia | 6 | <6 | 1755.1 | - |
| Abnormal liver enzyme | <6 | <6 | - | - |
| Musculoskeletal pain | 49 | 14 | 10129.3 | 7764.8 |
| Fatigue and malaise | 23 | 9 | 4611.6 | 4847.9 |
| Myositis | <6 | <6 | - | - |
| Myocarditis | <6 | <6 | - | - |
| Changes in the taste and smell | <6 | <6 | - | - |
| Generalized pain | 16 | <6 | 3508.5 | - |
| Fluid and electrolyte | <6 | <6 | - | - |
| Hair loss | 8 | <6 | 1614.9 | - |
| Chest pain | 19 | 7 | 3925.2 | 3894.7 |
| Fever and chills | 119 | 63 | 40369.2 | 52795.8 |
| Headache | 21 | 11 | 5367.9 | 7349.7 |
| Respiratory signs and symptoms | 313 | 150 | 64139.5 | 81570.8 |

**Figure S8**. Risks of incident PASC outcomes compared with the first infection cohort, by race/ethnicity subgroup. The error bars showed the 95% confidence interval (CI) of the estimated RR.

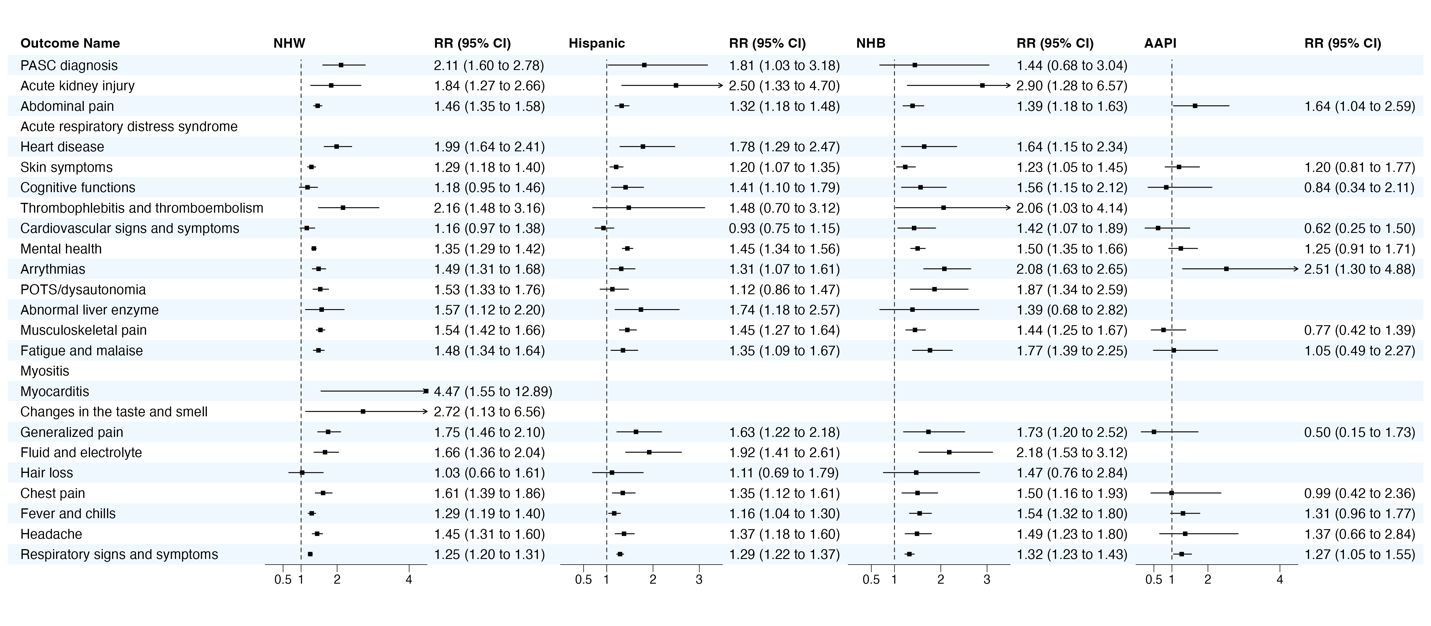

### Section S8 Supplemental Results: stratified analysis by obesity status subgroups

We conducted subgroup analysis by obesity status at cohort entry.

**Table S13.** Number of events and incidence rate per million persons per 6 months of patients in first and second infection episodes, after exact matching and propensity score matching, obese.

|  | Number of events | | Incidence rate per million persons per 6 months | |
| --- | --- | --- | --- | --- |
|  | First infection | Second infection | First infection | Second infection |
| PASC Diagnosis | 88 | 59 | 1027.9 | 2150.8 |
| Acute kidney injury | 31 | 16 | 434.7 | 699.3 |
| Abdominal pain | 1622 | 715 | 19433.6 | 26610.0 |
| Acute respiratory distress syndrome | <6 | <6 | - | - |
| Heart disease | 117 | 84 | 1371.3 | 3075.1 |
| Skin symptoms | 1375 | 544 | 17379.6 | 21366.1 |
| Cognitive functions | 271 | 112 | 3361.8 | 4326.6 |
| Thrombophlebitis and thromboembolism | 34 | 21 | 400.1 | 770.3 |
| Cardiovascular signs and symptoms | 446 | 162 | 5582.6 | 6362.0 |
| Mental health | 4226 | 1720 | 50804.1 | 64496.1 |
| Arrythmias | 426 | 206 | 5352.4 | 8074.2 |
| POTS/dysautonomia | 377 | 167 | 6104.0 | 8302.7 |
| Abnormal liver enzyme | 91 | 44 | 1111.1 | 1675.6 |
| Musculoskeletal pain | 1604 | 691 | 20005.7 | 26906.4 |
| Fatigue and malaise | 640 | 286 | 7524.1 | 10494.2 |
| Myositis | 8 | <6 | 108.4 | - |
| Myocarditis | <6 | 6 | - | 225.1 |
| Changes in the taste and smell | 8 | 9 | 133.2 | 462.1 |
| Generalized pain | 288 | 144 | 4173.4 | 6475.3 |
| Fluid and electrolyte | 122 | 60 | 1426.2 | 2188.4 |
| Hair loss | 97 | 35 | 1135.5 | 1279.7 |
| Chest pain | 515 | 269 | 6341.5 | 10357.4 |
| Fever and chills | 1155 | 537 | 23213.5 | 33004.5 |
| Headache | 1095 | 518 | 15744.6 | 23041.8 |
| Respiratory signs and symptoms | 5115 | 2172 | 60491.5 | 80037.5 |

**Table S14.** Number of events and incidence rate per million persons per 6 months of patients in first and second infection episodes, after exact matching and propensity score matching, non-obese.

|  | Number of events | | Incidence rate per million persons per 6 months | |
| --- | --- | --- | --- | --- |
|  | First infection | Second infection | First infection | Second infection |
| PASC Diagnosis | 46 | 39 | 601.3 | 1616.8 |
| Acute kidney injury | 40 | 21 | 594.5 | 991.1 |
| Abdominal pain | 1015 | 462 | 13470.0 | 19447.5 |
| Acute respiratory distress syndrome | 6 | <6 | 79.3 | - |
| Heart disease | 190 | 91 | 2484.6 | 3774.0 |
| Skin symptoms | 1361 | 571 | 19074.0 | 25398.2 |
| Cognitive functions | 282 | 138 | 3887.0 | 6024.1 |
| Thrombophlebitis and thromboembolism | 43 | 26 | 563.4 | 1081.0 |
| Cardiovascular signs and symptoms | 404 | 143 | 5541.7 | 6216.7 |
| Mental health | 2501 | 1170 | 33409.7 | 49325.4 |
| Arrythmias | 505 | 259 | 6999.1 | 11356.5 |
| POTS/dysautonomia | 265 | 140 | 4790.4 | 7695.7 |
| Abnormal liver enzyme | 54 | 32 | 735.9 | 1383.0 |
| Musculoskeletal pain | 889 | 418 | 12108.9 | 18033.2 |
| Fatigue and malaise | 558 | 264 | 7298.6 | 10988.3 |
| Myositis | <6 | <6 | - | - |
| Myocarditis | <6 | <6 | - | - |
| Changes in the taste and smell | <6 | <6 | - | - |
| Generalized pain | 160 | 95 | 2384.4 | 4485.9 |
| Fluid and electrolyte | 202 | 100 | 2640.5 | 4144.1 |
| Hair loss | 40 | 21 | 523.4 | 871.7 |
| Chest pain | 344 | 167 | 4650.8 | 7190.6 |
| Fever and chills | 1683 | 729 | 36372.4 | 47546.5 |
| Headache | 528 | 251 | 8302.6 | 12292.6 |
| Respiratory signs and symptoms | 5300 | 2122 | 69964.5 | 88876.6 |

**Figure S9**. Risks of incident PASC outcomes compared with the first infection cohort, by obesity status. The error bars showed the 95% confidence interval (CI) of the estimated RR.

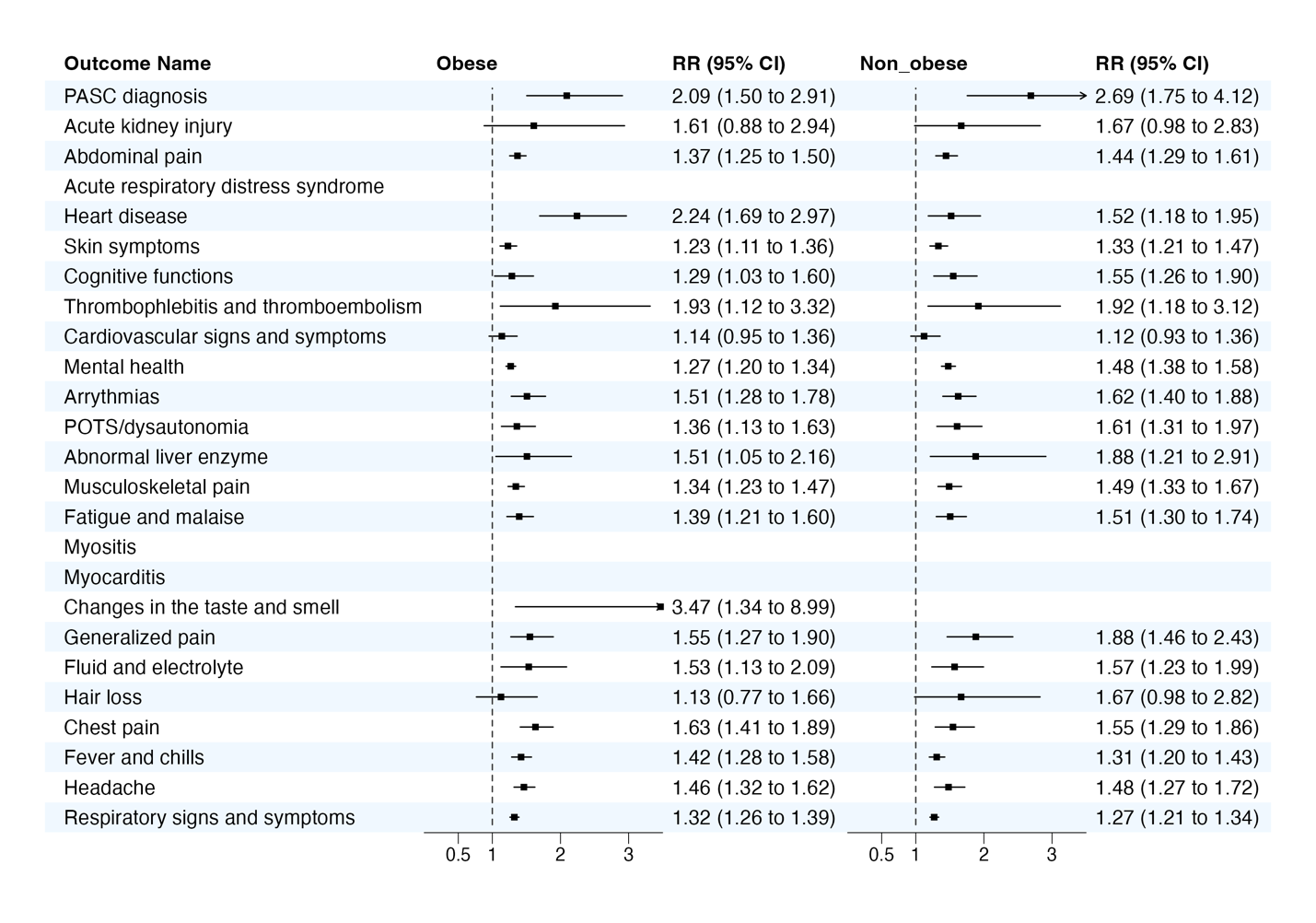

### Section S9 Supplemental Results: stratified analysis by severity of acute phase COVID-19 subgroups

We conducted subgroup analysis by the severity of acute phase COVID-19. The severity of COVID-19 at the index date was stratified into the following four levels: asymptomatic, mild (symptomatic but without severe complications), moderate (characterized by more serious conditions associated with COVID-19, such as gastroenteritis, dehydration, and pneumonia), and severe (marked by critical conditions necessitating ICU care or mechanical ventilation)^6^. For this sensitivity analysis, we grouped individuals presenting with either asymptomatic or mild symptoms into a “non-severe” category, while categorizing all remaining cases under a “severe” group.

**Table S15.** Number of events and incidence rate per million persons per 6 months of patients in first and second infection episodes, after exact matching and propensity score matching, severe.

|  | Number of events | | Incidence rate per million persons per 6 months | |
| --- | --- | --- | --- | --- |
|  | First infection | Second infection | First infection | Second infection |
| PASC Diagnosis | 17 | 7 | 1939.0 | 1449.0 |
| Acute kidney injury | 10 | 17 | 1339.8 | 4193.6 |
| Abdominal pain | 145 | 114 | 16995.0 | 24411.5 |
| Acute respiratory distress syndrome | <6 | <6 | - | - |
| Heart disease | 88 | 61 | 10111.7 | 12612.6 |
| Skin symptoms | 202 | 119 | 26198.1 | 28448.3 |
| Cognitive functions | 43 | 32 | 5233.6 | 7064.6 |
| Thrombophlebitis and thromboembolism | 34 | 23 | 3910.3 | 4788.4 |
| Cardiovascular signs and symptoms | 64 | 41 | 7707.0 | 8998.6 |
| Mental health | 381 | 337 | 44960.7 | 72055.0 |
| Arrythmias | 137 | 99 | 17414.7 | 23329.7 |
| POTS/dysautonomia | 46 | 28 | 7722.8 | 8296.1 |
| Abnormal liver enzyme | 23 | 18 | 2863.9 | 4132.9 |
| Musculoskeletal pain | 144 | 120 | 17124.7 | 25832.5 |
| Fatigue and malaise | 129 | 107 | 14806.3 | 22358.2 |
| Myositis | <6 | <6 | - | - |
| Myocarditis | <6 | <6 | - | - |
| Changes in the taste and smell | <6 | <6 | - | - |
| Generalized pain | 20 | 20 | 2693.0 | 4942.6 |
| Fluid and electrolyte | 92 | 74 | 10533.7 | 15388.6 |
| Hair loss | 7 | 9 | 799.8 | 1864.7 |
| Chest pain | 45 | 16 | 5367.5 | 3510.2 |
| Fever and chills | 212 | 133 | 45438.3 | 50609.3 |
| Headache | 59 | 42 | 8316.4 | 10786.4 |
| Respiratory signs and symptoms | 859 | 534 | 100628.4 | 114326.9 |

**Table S16.** Number of events and incidence rate per million persons per 6 months of patients in first and second infection episodes, after exact matching and propensity score matching, non-severe.

|  | Number of events | | Incidence rate per million persons per 6 months | |
| --- | --- | --- | --- | --- |
|  | First infection | Second infection | First infection | Second infection |
| PASC Diagnosis | 149 | 115 | 742.2 | 1863.6 |
| Acute kidney injury | 66 | 34 | 379.8 | 634.0 |
| Abdominal pain | 3216 | 1317 | 16185.1 | 21548.5 |
| Acute respiratory distress syndrome | 10 | <6 | 50.2 | - |
| Heart disease | 282 | 159 | 1403.8 | 2575.6 |
| Skin symptoms | 3216 | 1210 | 16919.0 | 20670.0 |
| Cognitive functions | 599 | 240 | 3135.3 | 4067.7 |
| Thrombophlebitis and thromboembolism | 55 | 31 | 274.3 | 503.0 |
| Cardiovascular signs and symptoms | 900 | 319 | 4733.5 | 5456.9 |
| Mental health | 7977 | 3247 | 40616.2 | 53656.4 |
| Arrythmias | 998 | 470 | 5270.7 | 8032.8 |
| POTS/dysautonomia | 788 | 358 | 5158.8 | 7466.4 |
| Abnormal liver enzyme | 189 | 76 | 964.3 | 1258.9 |
| Musculoskeletal pain | 3089 | 1354 | 16030.8 | 22798.5 |
| Fatigue and malaise | 1381 | 612 | 6919.0 | 9954.1 |
| Myositis | 14 | 6 | 77.4 | 107.7 |
| Myocarditis | <6 | 6 | - | 98.6 |
| Changes in the taste and smell | 13 | 14 | 84.1 | 291.7 |
| Generalized pain | 489 | 257 | 2878.1 | 4900.0 |
| Fluid and electrolyte | 307 | 143 | 1528.6 | 2317.2 |
| Hair loss | 167 | 59 | 831.3 | 955.4 |
| Chest pain | 1021 | 496 | 5266.9 | 8339.2 |
| Fever and chills | 3484 | 1453 | 26868.3 | 35468.8 |
| Headache | 1962 | 881 | 11574.9 | 16729.9 |
| Respiratory signs and symptoms | 11579 | 4549 | 57699.5 | 73878.3 |

### Section S10 Supplemental Results: stratified analysis by vaccine status

We conducted subgroup analysis by the vaccine status.

**Table S17.** Number of events and incidence rate per million persons per 6 months of patients in first and second infection episodes, after exact matching and propensity score matching, vaccinated.

|  | Number of events | | Incidence rate per million persons per 6 months | |
| --- | --- | --- | --- | --- |
|  | First infection | Second infection | First infection | Second infection |
| PASC Diagnosis | 57 | 49 | 1145.2 | 2901.1 |
| Acute kidney injury | 33 | 20 | 795.5 | 1429.4 |
| Abdominal pain | 1096 | 515 | 22492.2 | 31065.2 |
| Acute respiratory distress syndrome | <6 | <6 | - | - |
| Heart disease | 102 | 55 | 2057.6 | 3271.8 |
| Skin symptoms | 672 | 300 | 14443.5 | 19055.0 |
| Cognitive functions | 74 | 34 | 1549.0 | 2095.5 |
| Thrombophlebitis and thromboembolism | 28 | 9 | 564.7 | 534.7 |
| Cardiovascular signs and symptoms | 260 | 97 | 5624.2 | 6163.0 |
| Mental health | 3525 | 1504 | 72166.9 | 90703.4 |
| Arrythmias | 343 | 141 | 7620.8 | 9116.6 |
| POTS/dysautonomia | 422 | 189 | 9794.7 | 12990.9 |
| Abnormal liver enzyme | 90 | 45 | 1858.1 | 2748.5 |
| Musculoskeletal pain | 1180 | 526 | 25527.4 | 33245.1 |
| Fatigue and malaise | 549 | 258 | 11151.7 | 15407.6 |
| Myositis | 7 | <6 | 165.3 | - |
| Myocarditis | <6 | <6 | - | - |
| Changes in the taste and smell | 7 | <6 | 211.8 | - |
| Generalized pain | 206 | 91 | 5345.3 | 6829.4 |
| Fluid and electrolyte | 105 | 51 | 2115.7 | 3026.3 |
| Hair loss | 79 | 22 | 1591.8 | 1305.6 |
| Chest pain | 346 | 171 | 7474.6 | 10765.3 |
| Fever and chills | 379 | 206 | 10592.0 | 16757.1 |
| Headache | 757 | 387 | 17704.0 | 26403.0 |
| Respiratory signs and symptoms | 1840 | 862 | 37347.3 | 51585.5 |

**Table S18.** Number of events and incidence rate per million persons per 6 months of patients in first and second infection episodes, after exact matching and propensity score matching, unvaccinated.

|  | Number of events | | Incidence rate per million persons per 6 months | |
| --- | --- | --- | --- | --- |
|  | First infection | Second infection | First infection | Second infection |
| PASC Diagnosis | 142 | 94 | 764.2 | 1690.6 |
| Acute kidney injury | 81 | 55 | 497.1 | 1121.9 |
| Abdominal pain | 2867 | 1160 | 15670.5 | 21135.7 |
| Acute respiratory distress syndrome | 10 | 8 | 54.5 | 145.6 |
| Heart disease | 406 | 223 | 2185.2 | 4012.8 |
| Skin symptoms | 3188 | 1196 | 18221.4 | 22877.1 |
| Cognitive functions | 643 | 270 | 3646.9 | 5088.3 |
| Thrombophlebitis and thromboembolism | 83 | 63 | 448.2 | 1136.9 |
| Cardiovascular signs and symptoms | 898 | 298 | 5085.8 | 5627.6 |
| Mental health | 6509 | 2650 | 35878.1 | 48725.2 |
| Arrythmias | 1092 | 512 | 6208.1 | 9718.1 |
| POTS/dysautonomia | 605 | 261 | 4427.1 | 6219.2 |
| Abnormal liver enzyme | 158 | 69 | 879.6 | 1282.1 |
| Musculoskeletal pain | 2642 | 1169 | 14771.9 | 21778.9 |
| Fatigue and malaise | 1238 | 583 | 6703.7 | 10548.5 |
| Myositis | 11 | 6 | 64.8 | 117.5 |
| Myocarditis | 9 | 7 | 49.7 | 128.8 |
| Changes in the taste and smell | 9 | 11 | 60.7 | 246.0 |
| Generalized pain | 426 | 215 | 2637.3 | 4415.2 |
| Fluid and electrolyte | 390 | 218 | 2099.6 | 3924.2 |
| Hair loss | 138 | 42 | 743.5 | 756.4 |
| Chest pain | 913 | 438 | 5069.1 | 8134.0 |
| Fever and chills | 3839 | 1522 | 33357.9 | 42905.7 |
| Headache | 1645 | 686 | 10527.4 | 14513.8 |
| Respiratory signs and symptoms | 12239 | 4762 | 66248.9 | 86078.8 |
